# Patterns of analgesic prescribing and high-risk prescribing in primary care in Ireland 2014-2022 – a repeated cross-sectional study

**DOI:** 10.1101/2025.04.08.25325447

**Authors:** Molly Mattsson, Ahmed Hassan Ali, Fiona Boland, Michelle Flood, Ciara Kirke, Emma Wallace, Derek Corrigan, Mary E Walsh, Tom Fahey, Brian MacKenna, Frank Moriarty

**Affiliations:** School of Pharmacy and Biomolecular Sciences, Royal College of Surgeons in Ireland (RCSI) University of Medicine and Health Sciences, Dublin, Ireland; Data Science Centre, RCSI University of Medicine and Health Sciences, Dublin, Ireland; National Medication Safety Programme, HSE National Quality and Patient Safety Directorate, Dublin, Ireland; Department of General Practice, University College Cork, Cork, Ireland; Health Technology Assessment Directorate, Health Information and Quality Authority, Dublin, Ireland; School of Public Health, Physiotherapy and Sports Science, University College Dublin, Ireland; Department of General Practice, RCSI University of Medicine and Health Sciences, Dublin, Ireland; The Bennett Institute for Applied Data Science, Nuffield Department of Primary Care Health Sciences, University of Oxford, Oxford, United Kingdom

## Abstract

**Background:** Pain is a significant burden on individuals, healthcare systems, and society. Analgesic drugs carry many therapeutic benefits; however, all drugs are associated with adverse effects and risk of harm. Non-steroidal anti-inflammatory drugs (NSAIDs) and opioids have been identified as particularly high-risk due to the risk of side effects and/or dependency. This study aims to examine how patterns of analgesic prescribing have changed in primary care in Ireland between 2014 and 2022.

**Methods:** Monthly data on medicines prescribed and dispensed in primary care on the means-tested General Medical Services (GMS) scheme in Ireland was used. Prevalence, initiations, discontinuations, chronic use, and high-risk prescribing, as defined by Scottish Polypharmacy Guidance, were summarised per year.

**Results:** The prevalence of overall analgesic use decreased slightly over time, with 48.3% of GMS-eligible individuals dispensed an analgesic in 2014 and 46.3% in 2022. This was largely driven by decreasing NSAID use, from 29.4% in 2014 to 25.0% in 2022. Prevalence for all other analgesic drug classes increased, however after age/sex adjustment, higher odds of use in 2022 vs 2014 only persisted for gabapentinoids and amitriptyline. Some forms of high-risk prescribing increased over time, including NSAIDs dispensed with oral anticoagulants, corticosteroids, and SSRIs, with fewer decreasing.

**Discussion:** There was an overall reduction of analgesic use in Ireland, driven by decreasing systemic NSAID use. While most other analgesic drug classes are increasing, this may largely be explained by changing demographics, particularly the age profile of the population. Despite this, interventions addressing rising high-risk prescribing may be needed.

**Statement of Significance:** Analgesic drug classes are an important focus for improving medication safety. The findings of this study suggest an overall reduction of analgesic use in Ireland, driven by a decrease in systemic NSAID use. Increasing use of other analgesic drug classes may largely be explained by a change in demographics, particularly the age profile of the population. Analgesic use, and high-risk prescribing remains high and suggest a need for enhanced availability of and access to non-pharmacological services and interventions, as well as improved education and deprescribing support for healthcare professionals.

## Background

Pain is one of the most common reasons people seek medical care^1^. The prevalence of chronic pain estimated at 20-44% in different studies^2-4^. Pain management is a complex process influenced by clinical and patient-specific factors including diagnostic uncertainty, individual patient preferences, and challenges associated with managing both physical and mental health multimorbidities^5, 6^. Pharmacological management of pain depends on the type and duration of pain experienced. While drug treatments may offer many therapeutic benefits, all drugs have potential for adverse effects and harm, and some pose risk of dependency. Harm associated with acute pain treatment is more likely to be short-term or reversible, however prolonged analgesic use for chronic pain may present a variety of risks requiring careful consideration.

Among the risks of analgesic treatments, opioids are particularly high-risk and are no longer recommended as a first-line treatment for any form of chronic pain. In Ireland they are not routinely recommended for most cases of acute pain except in in-patient settings for short-term use as part of balanced multimodal analgesia^7^. Despite these guidelines, recent research in Ireland has shown an increase in opioid prescribing among individuals with public health cover rising from 14.4% in 2010 to 16.3% in 2019, with the greatest increase in the ≥65 years age group^8^. However, trends in opioid prescribing should be considered in the context of overall analgesic prescribing patterns. Changes may reflect variation in prevalence of pain conditions and the balance between medication initiation, chronic use and discontinuation. Decreased opioid use may lead to greater use of alternative analgesics, each with their own risks. For example, non-steroidal anti-inflammatory drugs (NSAIDs) are another important focus for medication safety, given increased risks of gastrointestinal bleeding, acute kidney injury, and adverse cardiovascular events, particularly in older adults^9^. Similarly gabapentinoid prescribing has increased globally since they were initially introduced along with increased misuse and abuse^10^, and increased mortality when used in combination with opioids^11^.

Reduction of medication-related harm has become an increasing priority internationally, driven by the World Health Organisation’s (WHO) Third Global Patient Safety Challenge, Medication Without Harm^12^. One of the challenge’s three focus areas is higher-risk situations, which includes prescribing of specific medications like opioids and NSAIDs. Several other national/international initiatives to reduce medication harm with some focus on analgesics have been developed. Scotland’s Polypharmacy Guidance highlights indicators of potentially inappropriate prescribing, including multiple indicators relating to NSAIDs, opioids and gabapentinoidss^13^. Similarly, three of the eleven Organisation for Economic Co-operation and Development (OECD) quality indicators for prescribing in primary care relate to analgesic drugs^14^. In addition, more specific interventions to assess and improve higher-risk prescribing have been implemented in various settings. In the UK, the roll-out of the PINCER intervention has involved training pharmacists working in general practice to provide feedback, education, and support to GP practices, systematically identifying and proactively reviewing high-risk prescribing, including NSAID use ^15^.

Given the increasing global emphasis on reducing medication-related harm and the well-documented risks associated with analgesics, it is important to understand prescribing trends in different settings. Primary care plays a key role in pain management and accounts for the largest volume of analgesia prescribing, making changes in prescribing practices particularly important with patient safety implications. Therefore, this study aims to examine changes in analgesic prescribing in Irish primary care between 2014 and 2022. By identifying trends in prescribing practices, the research will provide evidence that can inform patient safety initiatives and policy decisions.

## Methods

This is a repeated cross-sectional study of medicines dispensed in primary care in Ireland from 2014 to 2022. The protocol for the overarching project has been previously published^16^. The study was approved by the RCSI University of Medicine and Health Sciences Research Ethics Committee (ref: REC202201015) and Health Service Executive (HSE) Reference Research Ethics Committee B (ref: RRECB1022FM).

### Population and data sources

This study includes individuals eligible for the General Medical Services (GMS) scheme which is based on age and income and covers approximately 32% of the population, typically those more socioeconomically deprived than the general population^17^. Data on GMS prescribing is held by the Health Service Executive (HSE) Primary Care Reimbursement Service (PCRS), which processes pharmacy claims for dispensed medications. The PCRS pharmacy claims database is a national source widely used in pharmaceutical policy and pharmacoepidemiology research in Ireland^17^.

Data for this study were obtained from the PCRS through an information request and provided to the research team under a data transfer agreement. Analgesics were identified using WHO Anatomical Therapeutic Chemical (ATC) Classification codes and include opioids, paracetamol, NSAIDs, topical analgesics, antimigraine preparations, gabapentinoids, and low-dose amitriptyline.

See Supplementary Table 1. Data were at the level of individual medications dispensed to a patient. For each item, information included anonymised identifiers for individuals, date of dispensing, sex and age group of the individual, drug ATC code, product name and strength, quantity dispensed, and cost. Anonymised identifiers were created by the PCRS for the purpose of data sharing and cannot be linked to actual individuals by the researchers or others. Defined daily doses were applied to relevant ATC codes based route of administration^18^.

### Outcomes

Outcomes were derived to capture patterns of prescribed analgesia utilisation, including:

- Age group and sex-specific volume of use
- Prevalence of use, both overall and by age group and sex
- Prevalence of initiations, discontinuations, and chronic use
- Prevalence of high-risk use (defined below).

All outcomes were calculated for each year from 2014 to 2022. Outcomes were derived for all drug classes. For opioids, outcomes were further calculated for each individual drug, as well as weak and strong opioids and long-acting opioid formulations as outlined in Supplementary Table 1. Systemic NSAIDs, outcomes were also derived for COX-2 inhibitors and non-selective NSAIDs, and among topical products for lidocaine plasters, capsaicin, and topical NSAIDs. Products containing paracetamol were calculated for all paracetamol products, including opioid combinations, and non-combination products. Outcomes were also derived for other analgesics, specifically pregabalin, gabapentin, and low-dose amitriptyline.

Number of dispensings, cost, and standard doses were expressed as rates per 1,000 population by sex and age group. PCRS annual reports provide numbers of eligible individuals at the end of each calendar year^19^. Standard doses, accounting for prescription quantity, potency, pack size and strength, based on WHO Defined Daily Doses (DDD),^20^ were calculated for all medications. For opioids, oral morphine equivalents (OME) were also calculated. These were derived to both single and combination medications, with strengths mapped to standard dose for each component ingredient in the case of the latter (see Supplementary Table 2).

Pattern of use outcomes derived include prevalence of use, initiations, discontinuations, and chronic use, as well as high-risk polypharmacy dispensing indicators as defined by the Scottish Polypharmacy Guidance^13^. Prevalence, initiations, discontinuations, and chronic use were all further calculated by age group and sex. High-risk dispensing indicators related to NSAIDs dispensed with medications increasing the risk of bleeding, acute kidney injury, or seizures, opioids dispensed with medications increasing the risk of falls and delirium (outlined in Supplementary Table 3), and opioids and gabapentinoids dispensed at doses increasing the risk of dependency. Prevalence was calculated as the proportion of individuals with any occurrence, and percentage of dispensings of the relevant analgesic drug class considered high risk. An overview of the included outcomes is provided in Supplementary Table 4.

In addition to describe outcomes over time, the aim of the analysis was to explore trends in prevalence and determine whether a linear trend was present over time. To examine the prevalence of individual drug classes and the percentage of people with a high-risk dispensing, univariable and multivariable regression analyses were conducted, using a generalised linear model with a logit link and the binomial distribution. Univariable models included year as an independent variable, and multivariable models further adjusted for age group and sex. All analyses were conducted using Stata version 18 (College Station, TX: StataCorp LLC). Statistical significance was set at p<0.05.

## Results

The number of GMS eligible patients during the study period decreased from 1,768,700 in 2014 to 1,568,379 in 2022 (see Supplementary Table 5). Trends over time are reported descriptively as changes for the full GMS population. The prevalence of all analgesic use decreased slightly during the study period, with 48.3% of all GMS eligible individuals dispensed an analgesic in 2014 and 46.3% in 2022. The largest decrease in prevalence for a drug class was systemic NSAIDs, from 29.4% in 2014 to 25.0% in 2022. Prevalence for all other analgesic drug classes increased during the study period; opioids increased from 19.7% in 2014 to 20.8% in 2022, paracetamol from 29.7% in 2014 to 32.0% in 2022, topical analgesics from 14.0% to 14.9%, and other analgesics (low-dose amitriptyline, pregabalin, and gabapentin) from 7.6% in 2014 to 9.6% in 2022. For opioids, the individual drugs with the highest prevalence were codeine (which increased from 12.8% to 14.9%), tramadol (which decreased from 7.3% to 5.4%) and oxycodone (which increased from 1.2% to 2.1%), see Supplementary Table 6. All drug classes saw a decrease in prevalence of use in 2020, coinciding with the onset of the COVID-19 pandemic. Although prevalence increased again after 2020 for all drug classes, 2021 and 2022 prevalence were lower for all drug classes compared to 2019, with the exception of other analgesics (gabapentin, pregabalin, and low-dose amitriptyline). See Figure 1 and Supplementary Table 6.

**Figure 1.**
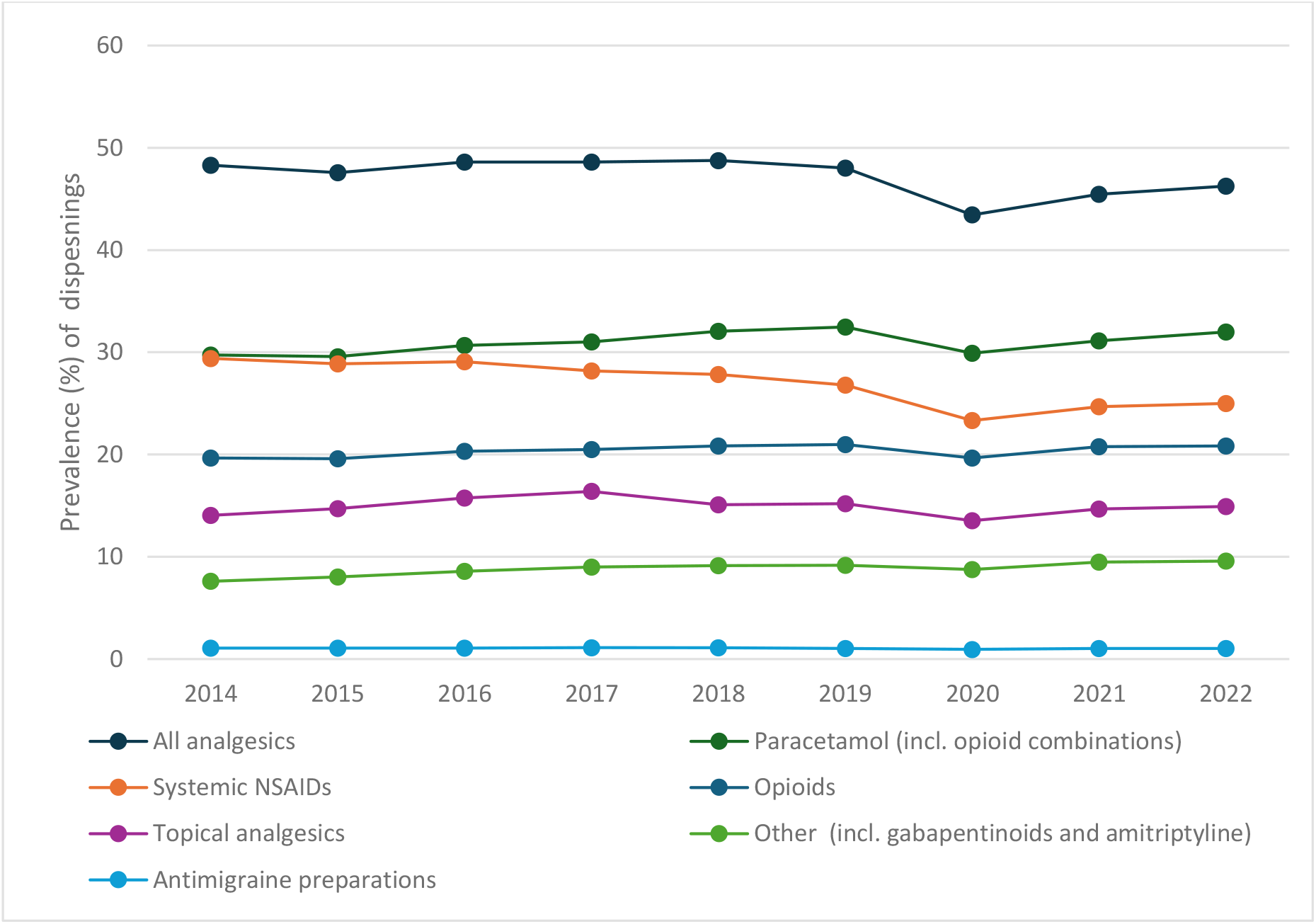
Prevalence of analgesic use between 2014 and 2022 by drug class.

For opioids, prevalence of use was consistently highest in the 75+ age group, remaining stable around 35%, followed by the 65-69, 55-64, and 70-74 age groups. Opioid use prevalence was stable throughout the study period for most age groups; however, decreases were seen for both 16-24 and 25-34 year olds, and an increase was seen for 65-69 year olds. Similar results were seen for gabapentinoids, with the highest prevalence seen in the oldest age groups. For systemic NSAIDs, prevalence decreased for all age groups during the study. The highest prevalence was amongst middle-aged adults, with the highest prevalences in those aged 45-54 years (39.2% in 2014 and 36.1% in 2022), followed by the 55-64, and 35-44 age groups. Comparatively lower prevalence was seen for the oldest age groups, with prevalence in those aged ≥75 decreasing from 28.8% to 22% (see Figure 2 and Supplementary Table 7).

**Figure 2.**
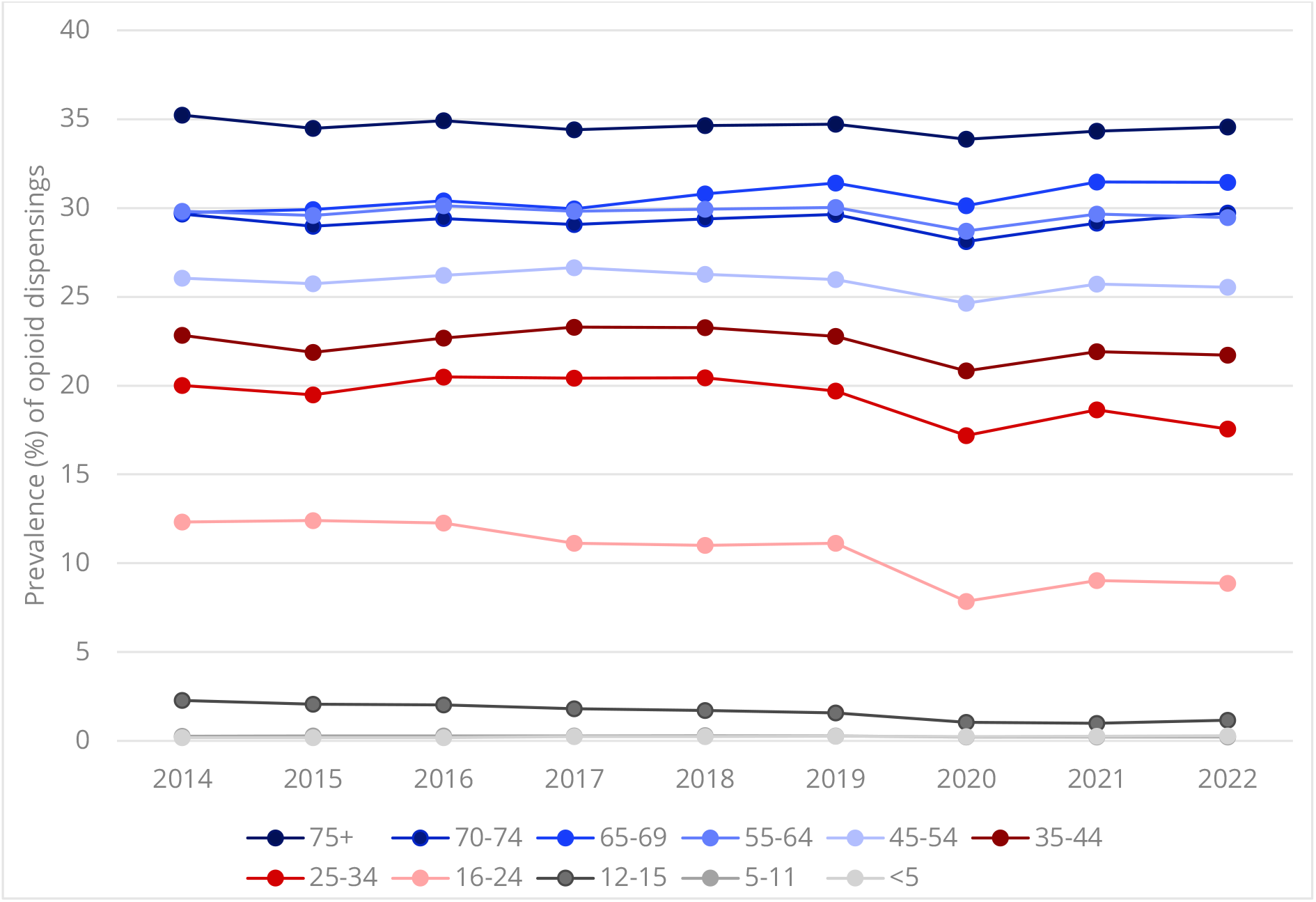
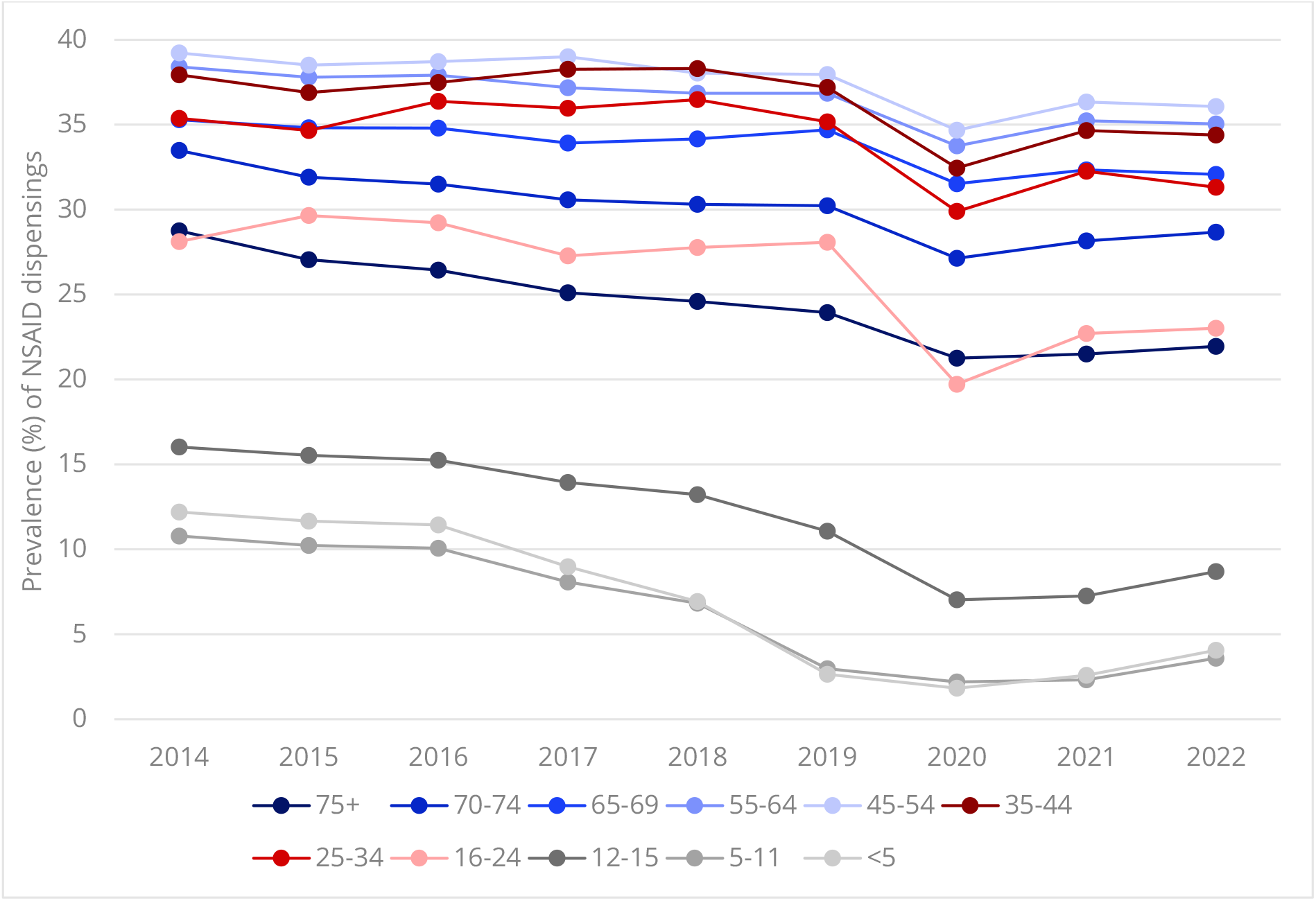
Prevalence of opioid (top) and systemic NSAID (bottom) use between 2014 and 2022 by age group.

Females had a higher prevalence of use for all drug classes; for opioids this was 23-24% during the study versus 16-17% in males. Prevalence of systemic NSAID use decreased from 33% to 29% in women, and from 25% to 20% in men (see Figure 3 and Supplementary Table 7). Rates of dispensings, cost, DDDs, and OMEs calculated by age group and sex followed a similar pattern to the prevalence of use. Full results are available as supplementary data (see data availability statement).

**Figure 3.**
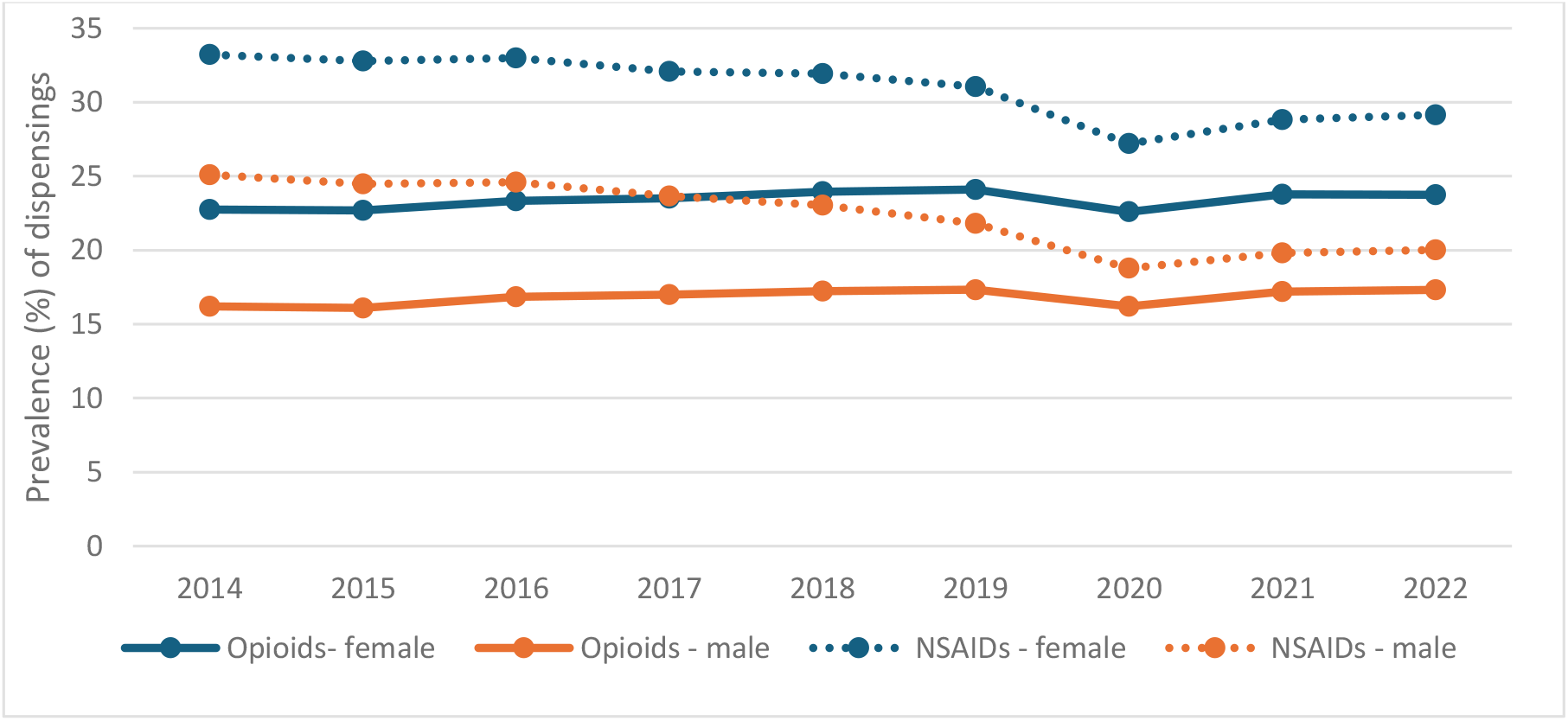
Prevalence of use of opioids (solid lines) and NSAIDs (dashed lines) for females (blue) and males (orange) between 2014 and 2022.

In regression models adjusting for age group and sex, the odds of being dispensed all drug classes decreased over the study, with the exception of other analgesics i.e. gabapentionoids and amitriptyline (see Table 1).

**Table 1.** Unadjusted and adjusted regression for the yearly change in prevalence of use (i.e. receipt of any dispensing) and high-risk dispensings

|  | Unadjusted OR | 95% CI | Adjusted<sup>2</sup> OR | 95% CI |
| --- | --- | --- | --- | --- |
| **Analgesic drug classes** |  |  |  |  |
| Analgesics | 0.98 | 0.98–0.98 | 0.98 | 0.98–0.98 |
| Opioids | 1.01 | 1.01–1.01 | 0.99 | 0.99–0.99 |
| Systemic NSAIDs | 0.96 | 0.96–0.96 | 0.96 | 0.96–0.96 |
| Paracetamol | 1.01 | 1.01–1.01 | 0.99 | 0.99–0.99 |
| Others<sup>1</sup> | 1.03 | 1.03–1.03 | 1.01 | 1.01–1.01 |
| Topical analgesics | 1 | 1.00–1.00 | 0.97 | 0.97–0.97 |
| Antimigraine | 0.99 | 0.99–0.99 | 0.99 | 0.99–0.99 |
| **High-risk medications/medication combinations** |  |  |  |  |
| NSAID with antiplatelet | 0.97 | 0.97–0.97 | 0.94 | 0.94–0.94 |
| NSAID with anticoagulant | 1.08 | 1.08–1.08 | 1.06 | 1.06–1.06 |
| NSAID with corticosteroid | 1.04 | 1.04–1.05 | 1.03 | 1.02–1.03 |
| NSAID with SSRI | 1.04 | 1.04–1.04 | 1.03 | 1.03–1.03 |
| NSAID with ACE/ARB and diuretic | 0.98 | 0.98–0.98 | 0.95 | 0.95–0.95 |
| NSAID with ACE/ARB and metformin (over 65s) | 0.91 | 0.91–0.92 | 0.91 | 0.91–0.92 |
| NSAID with lithium | 0.99 | 0.98–1.00 | 0.98 | 0.97–0.98 |
| Opioid with ≥2 other sedating or anticholinergic drugs (over 65s) | 0.98 | 0.98–0.99 | 0.98 | 0.98–0.98 |
| High-dose opioid | 1.02 | 1.02–1.03 | 1 | 1.00–1.00 |
| High-dose pregabalin | 1.01 | 1.01–1.01 | 1 | 1.00–1.01 |
| High-dose gabapentin | 1.04 | 1.03–1.05 | 1.02 | 1.01–1.03 |

<sup>1</sup>pregabalin, gabapentin, and low-dose amitriptyline
<sup>2</sup>adjusted for age group and sex

Between 2015 and 2022, initiations of opioids (12.7% versus 14.0%) and paracetamol (13.2% versus 14.5%) increased slightly (Table 2). The prevalence of initiations dropped in 2020 for all drug classes and remained below 2019 levels in 2021 and 2022. Discontinuations remained relatively stable for all drug classes over the study period, while there were increases in prevalence of chronic use of opioids (6.1% versus 7.5%), paracetamol (5.0% versus 7.7%), and other analgesics (4.7% and 6.2%). The percentage of all opioid dispensings constituting chronic use increased from approximately 30% to 35%, while the percentage of chronic paracetamol dispensings increased from 29% to 38% of all paracetamol dispensings (see Supplementary Table 6). Full results by age group and sex are available as supplementary data (see data availability statement, and absolute numbers of individuals are reported in Supplementary Table 8.

**Table 2.** Prevalence (%) of initiations, discontinuations, and chronic use of analgesic drug classes in the GMS population^**1**^

|  | 2015 | 2016 | 2017 | 2018 | 2019 | 2020 | 2021 | 2022 |
| --- | --- | --- | --- | --- | --- | --- | --- | --- |
| **Initiations** |  |  |  |  |  |  |  |  |
| Opioids | 12.7 | 13.7 | 14.0 | 14.2 | 14.3 | 13.2 | 13.9 | 14.0 |
| Systemic NSAIDs | 19.5 | 21.4 | 21.6 | 21.6 | 21.2 | 18.6 | 19.7 | 19.8 |
| Paracetamol<sup>2</sup> | 18.6 | 20.3 | 20.9 | 21.6 | 21.7 | 19.5 | 19.9 | 20.4 |
| Antimigraine preparations | 0.7 | 0.7 | 0.7 | 0.7 | 0.7 | 0.6 | 0.7 | 0.7 |
| Topical analgesics | 10.9 | 12.1 | 12.6 | 11.9 | 12.0 | 10.4 | 11.1 | 11.3 |
| Other analgesics<sup>3</sup> | 4.2 | 4.4 | 4.5 | 4.4 | 4.4 | 4.0 | 4.4 | 4.4 |
| **Discontinuations** |  |  |  |  |  |  |  |  |
| Opioids | 10.5 | 11.3 | 11.6 | 11.7 | 11.8 | 10.7 | 11.3 | 11.5 |
| Systemic NSAIDs | 14.6 | 15.9 | 16.2 | 16.2 | 16.4 | 14.5 | 15.3 | 15.5 |
| Paracetamol<sup>2</sup> | 14.3 | 15.5 | 15.9 | 16.0 | 16.1 | 14.5 | 14.8 | 15.1 |
| Antimigraine preparations | 0.5 | 0.6 | 0.6 | 0.6 | 0.6 | 0.5 | 0.5 | 0.5 |
| Topical analgesics | 9.4 | 10.5 | 11.3 | 11.0 | 10.6 | 9.2 | 9.6 | 10.0 |
| Other analgesics<sup>3</sup> | 3.3 | 3.7 | 3.8 | 3.8 | 3.7 | 3.3 | 3.6 | 3.6 |
| **Chronic use** |  |  |  |  |  |  |  |  |
| Opioids | 6.1 | 6.4 | 6.6 | 6.9 | 7.2 | 7.3 | 7.7 | 7.5 |
| Systemic NSAIDs | 3.6 | 3.5 | 3.5 | 3.5 | 3.5 | 3.6 | 3.8 | 3.6 |
| Paracetamol<sup>2</sup> | 8.5 | 9.0 | 9.4 | 10.1 | 10.6 | 11.2 | 12.1 | 12.2 |
| Antimigraine preparations | 0.3 | 0.3 | 0.3 | 0.4 | 0.3 | 0.3 | 0.3 | 0.4 |
| Topical analgesics | 2.9 | 3.3 | 3.5 | 2.8 | 3.0 | 3.3 | 3.8 | 3.8 |
| Other analgesics<sup>3</sup> | 4.7 | 5.1 | 5.4 | 5.6 | 5.8 | 5.7 | 6.1 | 6.2 |

<sup>1</sup>Initiations and discontinuations are defined based on a medication-free interval of 90 days before the first dispensing and after the last dispensing respectively, and chronic use for >90 days
<sup>2</sup>Including opioid combinations
<sup>3</sup>Low-dose amitriptyline, gabapentin, pregabalin

The prevalence of high-risk dispensings varied during the study period. All high-risk NSAID dispensings relating the risk of bleeding increased, with the exception of non-selective NSAIDs (dispensed with antiplatelets. The largest changes were seen for non-selective NSAIDs dispensed with SSRIs and opioids dispensed with sedating agents in those aged ≥65 years. Among those receiving NSAIDs, the percentage also dispensed an SSRI increased from 3.0% to 4.2%. The percentage of those aged ≥65 years receiving an opioid dispensing with two or more sedating or anticholinergic drugs decreased from 15.4% in 2014 to 14.0% in 2022. Indicators relating to NSAIDs and risk of acute kidney injury decreased during the study period. See Supplementary Table 9 and Figure 4. Full results by age group and sex are available as supplementary data (see data availability statement). In regression analysis, the change over time in the odds of having a high-risk dispensing was similar after adjusting for age group and sex (Table 1).

**Figure 4.**
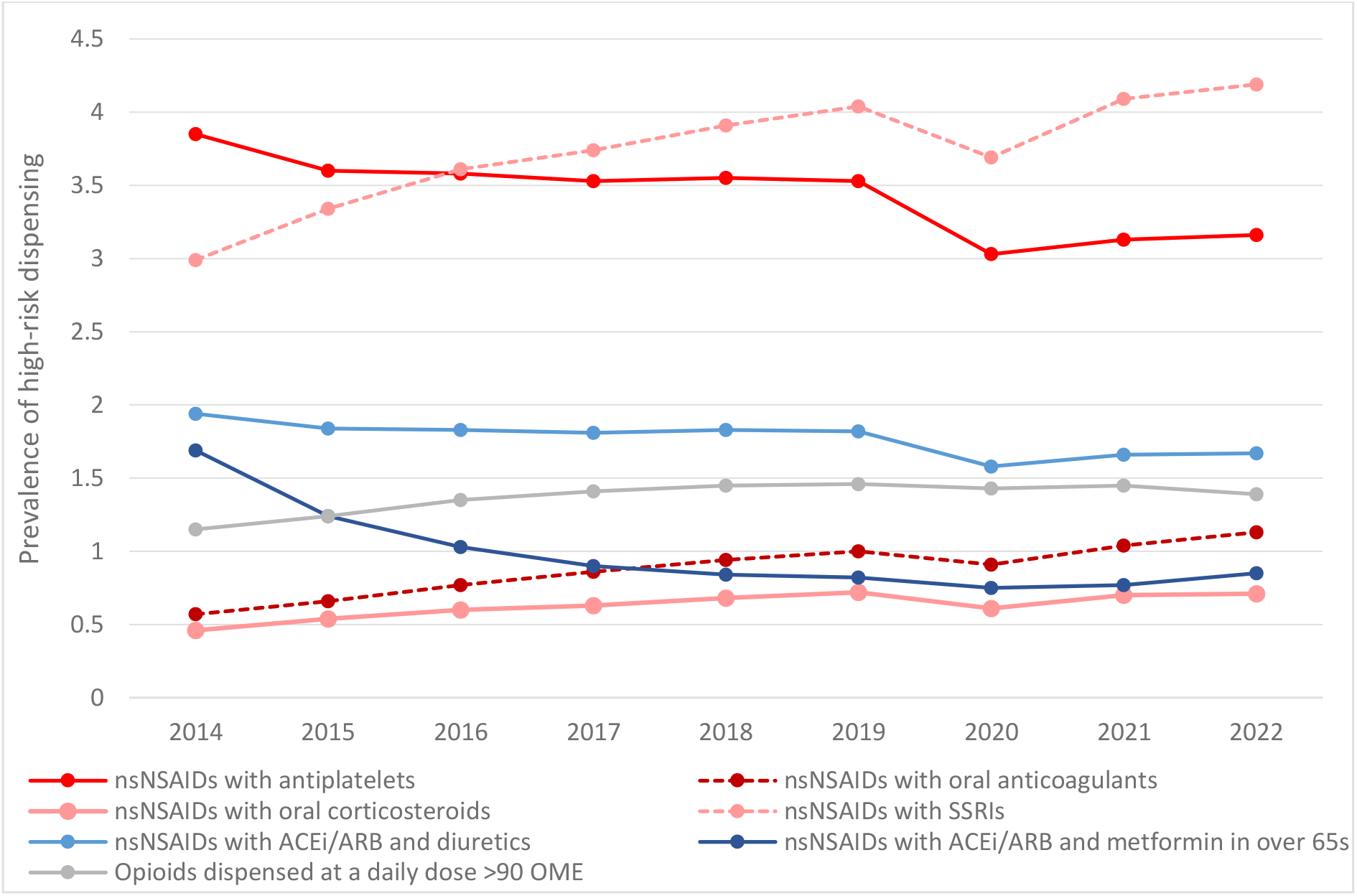
Prevalence of high-risk dispensing among users of the relevant analgesic drug class relating to bleeding risk (red), acute kidney injury risk (blue) and dependency risk (grey) * Indicators with prevalence <0.4% or >13% not shown.

## Discussion

The prevalence of analgesic use in Ireland across most drug classes increased between 2014 and 2022, including opioids, gabapentinoids, and paracetamol. However, this increase was offset by a decrease in NSAID use, and overall the prevalence of analgesia use decreased slightly. This is consistent with previous research on analgesia prescribing in Ireland^8, 21^. Previous research has identified higherrates of analgesia dispensings in Ireland compared to England^22^. The results of this study indicate higher prevalence of opioid use compared to Australia^23^. Additionally, the overall prevalence of analgesic use in Ireland appears higher than in Germany, the UK, and France^24^. However, the regression results, accounting for age-group and sex changes in the population over the study period, indicate a lower chance over time of an individual being prescribed any analgesic class, with the exception of gabapentinoids and low-dose amitriptyline. Importantly, the prevalence of several forms of high-risk dispensing is rising, independent of demographic changes, particularly those relating to bleeding risk with NSAIDs.

All analgesic drug classes saw a decrease in prevalence of use in 2020, coinciding with the COVID-19 pandemic. Although this increased since, 2021 and 2022 prevalence remained lower than in 2019 for all drug classes bar gabapentinoids and amitriptyline. Previous research in England found that the ‘lockdown’ measures accompanying the COVID-19 pandemic did not impact opioid prescribing in primary care^25^, with similar results identified in the Netherlands^26^. Canadian research on the experience of patients with chronic pain during the pandemic suggested a negative impact on patients’ experience of their care, with changes in treatment impacted by lack of access to prescribers/cancellation of medical appointments, and increased medication prescribing in compensation for stopping physical/psychological treatments^27^. A similar increase was not seen in Ireland, and in fact the observed decrease in 2020 could suggest the pandemic impacted access to prescribed analgesic drugs.

The prevalence of opioid, NSAID, and gabapentinoid use were all persistently higher in women compared to men. It is well-established that women have a higher prevalence of chronic pain and are prescribed more analgesics compared to men, although the mechanisms behind this discrepancy are not fully understood^28^. The highest prevalence of opioid and gabapentinoid use was consistently seen in the oldest age groups. Middle-aged groups had the highest prevalence of systemic NSAID use, with lower prevalence among the oldest groups. NSAIDs in older adults have been identified as high-risk prescribing, given renal and cardiovascular harms^29^, and the lower prevalence in these age groups is therefore a positive sign.

The prevalence of high-risk opioid dispensing with two or more sedatives in those aged ≥65 years decreased during the study period, which may suggest a move away from inappropriate polypharmacy in this vulnerable age group. The prevalence of NSAIDs dispensed with an SSRI increased, which may be a result of the global increase in SSRI-prescribing in recent years^30-32^, a trend which has also been seen in Ireland^33^. Conversely, NSAIDs dispensed with drugs increasing the risk of acute kidney injury decreased slightly. In recent years there have been efforts made to reduce this type of high-risk prescribing. In the UK, the rollout of the pharmacist-led PINCER intervention, targeting patients at risk of medication-harm, across 343 general practices reduced hazardous prescribing by 16.7% at six months and 15.3% at 12 months postintervention^34^. Similarly, the Data-Driven Quality Improvement in Primary Care (DQIP) randomised controlled trial in Scotland found that an intervention, comprising professional education, informatics, and financial incentives to review patients’ records to assess appropriateness, significantly reduced targeted high-risk prescribing^35^. The iSIMPATHY project, introduced pharmacists in Scotland, Northern Ireland, and Ireland to conduct medication reviews^36^. In Ireland, 70.7% of the polypharmacy indicator occurrences identified were resolved post review,^37^ suggesting structured medicines review are an important strategy to consider for improving quality of prescribing.

Ireland has historically had comparatively high rates of inappropriate prescribing, with one study comparing prescribing in 45-64 year olds in the GMS population in Ireland and the general population in Northern Ireland finding the prevalence in Ireland of potentially inappropriate prescribing more than double that in Northern Ireland^38^. Recent research has shown an overall reduction in high-risk prescribing in Ireland, with strong associations with age and number of chronic medications and with significant variation between prescribers^39^. Greater availability of prescribing data in Ireland would enable audit and feedback interventions for quality improvement of prescribing in these areas. Previous educational interventions involving audit and feedback to improve the quality of prescribing had positive results in Ireland, e.g. the Red-Green antibiotics initiative aimed at community prescribers, which saw an overall increase of 4.3% in prescribing of recommended antibiotics in the first 15 months of the initiative^40^.

This study includes individuals eligible for the GMS scheme, which is means-tested and covers approximately 32% of the population. Previous research has identified a higher prevalence of pain among older adults and individuals from disadvantaged backgrounds^3^. A recent study on opioid prescribing in England found a strong association of socioeconomic deprivation with opioid prescribing in primary care^41^. In addition to deprivation being linked to higher analgesic prescribing, it is also linked to developing multimorbidity earlier in life and a more rapid accumulation of physical and mental health conditions^42^, which due to treatment complexity and the single disease focus of clinical guidelines underpinning care may contribute to high-risk prescribing and inappropriate polypharmacy^43^. In that context, it is therefore likely that the Irish GMS population has higher prevalence of analgesia use compared to the general population.

Waiting times for GMS patients to access specialist care are extensive, resulting in patients with severe degenerative related chronic pain potentially waiting several years for joint replacement surgery, thus requiring strong analgesics in the meantime. In January 2025, over 61,000 individuals were awaiting an initial orthopaedic out-patient appointment and nearly 12,000 an orthopaedic inpatient appointment (including joint replacement procedures), with around 7,500 and 1,000 people respectively waiting for more than 12 months^44^. Additionally, access to non-pharmacological interventions, including exercise programmes, psychological therapies, acupuncture, and other treatments (e.g. transcutaneous electrical nerve stimulation) for GMS patients is often limited, despite some or all of these interventions being routinely recommended for multiple types of pain (e.g. osteoarthritis, rheumatoid arthritis, sciatica)^45-47^. For chronic primary pain (i.e. pain with no underlying condition adequately accounting for the pain or its impact), National Institute for Health and Care Excellence (NICE) guidelines recommend non-pharmacological interventions as primary interventions, with the use of opioids, gabapentinoids, paracetamol, and NSAIDs all discouraged^48^. Although these recommendations have come under criticism for their restrictiveness^49^, access to these non-pharmacological interventions is vital.

The main strength of this study is the use of comprehensive data, which has high accuracy and completeness given its primary purpose for pharmacy reimbursement. Further, dispensing-level data linked to individuals permitted examination of patterns of analgesic in greater detail than is possible with aggregate data. A limitation is the study only includes data relating to the means-tested GMS eligible population, with the older and more disadvantaged population not fully representative of the general population. Secondly, the data lacks details of indication for the prescription and patients’ diagnoses, so although drug classes examined are almost exclusively used for pain, we could not examine medication use by type of pain. Although all dispensings related to prescriptions issued in primary care, some of these medications may have been originally initiated in specialist settings. Finally, as we do not have access to eligibility start or end dates, or data on mortality, we are not eligible to account for these factors when analysing initiation and discontinuation patterns, and as data used relates to dispensings, we do not have information on individuals’ adherence to dispensed medicines.

## Conclusions

The findings of this study suggest an overall reduction of analgesic use in Ireland, driven by a decrease in use of systemic NSAIDs. Although prevalence of use for most other drug classes is increasing, this may largely be explained by the increasingly older age profile of the population. Despite improvements in some high-risk indicators, prescribing which increases risk of bleeding and chronic use of some drug classes remains a concern, and suggests a need for effective medicines review services, enhanced availability of non-pharmacological interventions, as well as measures to reduce waiting times for specialist care. It is important that further meaningful analgesic drug utilisation research is enabled, through timely access to appropriate data, in order to support policies and practices to ultimately improve medication appropriateness and patient safety.

## Acknowledgements

We would like to acknowledge the wider Controlled Drug Prescribing (CDRx) team.

## Data availability

The datasets analysed during the current study are not publicly available as this was not covered by the Data Exchange Agreement between RCSI and HSE-PCRS. Data can be requested from HSE-PCRS via an information request as detailed at https://www.hse.ie/eng/staff/pcrs/pcrs-publications/. Full results for initiations, discontinuations, chronic use, and high-risk dispensings by age group and sex are available from Zenodo (doi: 10.5281/zenodo.14870683)

## Author contributions

FM conceived the study. All authors were involved in the design of the study. MM and FM collected and curated the data. MM, FM, AHA developed the analysis plan. MM and FM conducted the analysis of the data. All authors were involved the interpretation of the analysis results. MM drafted the manuscript, and MF, AHA, EW, FB, CK, MEW, DC, TF, BMacK, and FM critically revised the manuscript. FM acquired funding for the study.

## Table legends

Table 1 Unadjusted and adjusted regression for the yearly change in prevalence of use and high-risk dispensings

Table 2. Prevalence (%) of initiations, discontinuations, and chronic use of analgesic drug classes in the GMS population

Table 3. Prevalence (%) of high risk dispensings in the GMS population

## Figure legends

Figure 1. Prevalence of use by drug class

Figure 2a. Prevalence of opioid use by age group

Figure 2b. Prevalence of systemic NSAID use by age group

Figure 3 Prevalence of use of opioids (solid lines) and NSAIDs (dashed lines) for females (blue) and males (orange)

Figure 4. Percentage of dispensings of the relevant analgesic drug class considered high risk

## Notes

### Competing Interest Statement

The authors have declared no competing interest.

### Funding Statement

This study is funded by the Health Research Board in Ireland (HRB) through the Secondary Data Analysis Projects scheme (CDRx project, grant number SDAP-2019-023). The funder had no role in in study design; in the collection, analysis, and interpretation of data; in the writing of this paper; or in the decision to submit this paper for publication. EW is funded by an HRB Emerging Clinician Scientist Award (grant number: ECSA/2020/002). MEW is funded by an HRB Applying Research into Policy and Practice Award (ARPP/2020/004).

### Author Declarations

The study was approved by the RCSI University of Medicine and Health Sciences Research Ethics Committee (ref: REC202201015) and Health Service Executive (HSE) Reference Research Ethics Committee B (ref: RRECB1022FM).

