## Supplementary for "Patterns of analgesic prescribing and high-risk prescribing in primary care in Ireland 2014-2022 – a repeated cross-sectional study"

**Supplementary table 1. Drugs included in the analysis**

| **Medication** | **ATC-code** |
| --- | --- |
| **Opioids** |  |
| ***Strong opioids (as defined by BNF)^1^*** |  |
| Morphine, incl. combinations^2^ | N02AA01, N02AA51 |
| Hydromorphone^2^ | N02AA03 |
| Oxycodone, incl. combinations^2^ | N02AA05, N02AA55 |
| Pethidine | N02AB02 |
| Fentanyl^3^ | N02AB03 |
| Buprenorphine^3^ | N02AE01 |
| Tramadol^2^ | N02AX02, N02AJ13, N02AJ14 |
| Tapentadol^2^ | N02AX06 |
| ***Weak opioids*** |  |
| Meptazinol | N02AX05 |
| Dihydrocodeine, incl. combinations | N02AA08, N02AJ01 |
| Codeine, incl. combinations | R05DA04, N02AJ06 |
| **Systemic NSAIDs** |  |
| ***Coxibs*** |  |
| Celecoxib | M01AH01 |
| Etoricoxib | M01AH05 |
| ***Non-selective NSAIDs*** |  |
| Indometacin | M01AB01 |
| Diclofenac, incl. combinations | M01AB05, M01AB55 |
| Aceclofenac | M01AB16 |
| Meloxicam | M01AC06 |
| Ibuprofen | M01AE01 |
| Naproxen, incl. combinations | M01AE02, M01AE52 |
| Ketoprofen | M01AE03 |
| Flurbiprofen | M01AE09 |
| Dexketoprofen | M01AE17, N02AJ14 |
| Mefenamic acid | M01AG01 |
| Nabumetone | M01AX01 |
| **Paracetamol, incl. combinations** | N02BE01, N02BE51 |
| **Topical analgesics** |  |
| ***Topical NSAIDs*** |  |
| Benzydamine | M02AA05 |
| Etofenamate | M02AA06 |
| Piroxicam | M02AA07 |
| Ketoprofen | M02AA10 |
| Ibuprofen | M02AA13 |
| Diclofenac | M02AA15 |
| ***Other topical agents*** |  |
| Capsaicin | M02AB01 |
| Lidocaine | N01BB02 |
| **Other analgesics** |  |
| Gabapentin | N03AX12 |
| Pregabalin | N03AX16 |
| Amitriptyline 10mg | N06AA09 |
| **Antimigraine preparations** |  |
| Sumatriptan | N02CC01 |
| Naratriptan | N02CC02 |
| Zolmitriptan | N02CC03 |
| Rizatriptan | N02CC04 |
| Almotriptan | N02CC05 |
| Eletriptan | N02CC06 |
| Frovatriptan | N02CC07 |
| Clonidine | N02CX02 |

^1^Joint Formulary Committee. British national formulary 78. British Medical Association and Royal Pharmaceutical Society of Great Britain; 2024

^2^Including long-acting oral formulations

^3^Including long-acting transdermal patches

**Supplementary table 2. Oral morphine equivalents (OME)**

| **Drug** | **OME equivalent** | **Source** |
| --- | --- | --- |
| Morphine (oral) | 1 | Nielsen et al^1^ |
| Morphine (rectal) | 1 | Not available, assumed same as oral |
| Morphine (parenteral) | 3 | Nielsen et al |
| Hydromorphone (oral) | 5 | Nielsen et al |
| Hydromorphone (parenteral) | 17.5 | Nielsen et al |
| Oxycodone (oral) | 1.5 | Nielsen et al |
| Oxycodone (parenteral) | 3 | Nielsen et al |
| Dihydrocodeine | 0.1 | Nielsen et al |
| Pethidine (oral) | 0.1 | Nielsen et al |
| Pethidine (parenteral) | 0.4 | Nielsen et al |
| Fentanyl (oral) | 100 | Nielsen et al |
| Fentanyl (nasal) | 160 | Curtis et al^2^ |
| Fentanyl (transdermal) | 112.5 | Nielsen et al |
| Buprenorphine (oral) | 38.8 | Nielsen et al |
| Buprenorphine (parenteral) | 75 | Nielsen et al |
| Buprenorphine (transdermal) | 91.67 | Nielsen et al |
| Tramadol | 0.2 | Nielsen et al |
| Meptazinol | 0.03 | Curtis et al |
| Tapentadol | 0.4 | Nielsen et al |
| Codeine (oral) | 0.1 | Nielsen et al |
| Codeine (parenteral) | 0.3 | Nielsen et al |

1.	Nielsen S, Degenhardt L, Hoban B, Gisev N. A synthesis of oral morphine equivalents (OME) for opioid utilisation studies. *Pharmacoepidemiology and Drug Safety*. 2016;25(6):733-737. doi:<https://doi.org/10.1002/pds.3945>

2.	Curtis HJ, Croker R, Walker AJ, Richards GC, Quinlan J, Goldacre B. Opioid prescribing trends and geographical variation in England, 1998&#x2013;2018: a retrospective database study. *The Lancet Psychiatry*. 2019;6(2):140-150. doi:10.1016/S2215-0366(18)30471-1

**Supplementary table 3. Anticholinergic drugs^1^**

| **Score 1** | | **Score 2** | | **Score 3** | |
| --- | --- | --- | --- | --- | --- |
| **Drug** | **ATC** | **Drug** | **ATC** | **Drug** | **ATC** |
| Alimemazine | R06AD01 | Amantadine | N04BB01 | Amitriptyline | N06AA09 |
| Alverine | A03AX08, A03AX58 | Belladonna alkaloids | A03BA01, A03BA04,  A03BB01, A03BB02,  A03BB03, A03BB04,  A03BB05, A03BB06,  A06AB30 | Amoxapine | N06AA17 |
| Alprazolam | N05BA12 | Carbamazepine | N03AF01 | Atropine | A03BA01, A03CB03 |
| Atenolol | C07AB03, C07FB03,  C07CB03, C07CB53,  C07BB03, C07DB01 | Cyclobenzaprine | M03BX08 | Benzatropine | N04AC01 |
| Brompheniramine maleate | R06AB01, R06AB51 | Cyproheptadine | R06AX02 | Brompheniramine | R06AB01, R06AB51 |
| Bupropion hydrochloride | N06AX12, A08AA62 | Levomepromazine | N05AA02 | Carbinoxamine | R06AA08 |
| Captopri | C09AA01, C09BA01 | Loxapine | N05AH01 | Chlorphenamine | R06AB04, R06AB54 |
| Chlortalidone | C03BA04, C03BB04,  C03EA06 | Molindone | N05AE02 | Chlorpromazine | N05AA01 |
| Cimetidine hydrochloride | A02BA01, A02BA51 | Oxcarbazepine | N03AF02 | Clemastine | R06AA04, R06AA54 |
| Clorazepate | N05BA05 | Pethidine hydrochloride | N02AB02, N02AG03,  N02AB52, N02AB72 | Clomipramine | N06AA04 |
| Codeine | R05DA04, N02AJ06,  N02AJ07, N02AJ08,  N02AJ09, N02AA59,  N02AA79, | Pimozide | N05AG02 | Clozapine | N05AH02 |
| Colchicine | M04AC01 |  |  | Darifenacin | G04BD10 |
| Diazepam | N05BA01 |  |  | Desipramine | N06AA01 |
| Digoxin | C01AA05 |  |  | Dicycloverine | A03AA07 |
| Dipyridamole | B01AC07 |  |  | Dimenhydrinate | R06AA11 |
| Disopyramide phosphate | C01BA03 |  |  | Diphenhydramine | R06AA02, R06AA52 |
| Fentanyl | N02AB03 |  |  | Doxepin | N06AA12 |
| Furosemide | C03CA01, C03CB01,  C03EB01 |  |  | Flavoxate | G04BD02 |
| Fluvoxamine | N06AB08 |  |  | Hydroxyzine | N05BB01,N05BB51 |
| Haloperidol | N05AD01 |  |  | Hyoscyamine | A03BA03, A03CB31 |
| Hydralazine | C02DB02, C02LG02 |  |  | Imipramine | N06AA02, N06AA03 |
| Hydrocortisone | H02AB09 |  |  | Mepyramine | R06AC01, R03DA12 |
| Isosorbide | C01DA08, C01DA58,  C05AE02, C01DA14 |  |  | Meclozine | R06AE05, R06AE55 |
| Loperamide | A07DA03, A07DA05,  A07DA53 |  |  | Nortriptyline | N06AA10 |
| Metoprolol | C07AB02, C07FX03,  C07FB13, C07FB02,  C07FX05, C07CB02,  C07BB02, C07BB52 |  |  | Olanzapine | N05AH03 |
| Morphine | N02AA01, N02AA51,  N02AG01, A07DA52,  R05DA05 |  |  | Orphenadrine | N04AB02, M03BC01,  M03BC51 |
| Nifedipine | C08CA05, C07FB03,  C08GA01, C08CA55 |  |  | Oxybutynin | G04BD04 |
| Prednisone | H02AB07, A07EA03 |  |  | Paroxetine | N06AB05 |
| Quinidine | C01BA01, C01BA51,  C01BA71 |  |  | Perphenazine | N05AB03 |
| Ranitidine | A02BA02, A02BA07 |  |  | Procyclidine | N04AA04 |
| Risperidone | N05AX08 |  |  | Promazine | N05AA03 |
| Theophylline | R03DA04, R03DB04,  R03DA54, R03DA74 |  |  | Promethazine | R06AD02, R06AD52, |
| Trazodone | N06AX05 |  |  | Propantheline | A03AB05, A03CA34 |
| Triamterene | C03DB02 |  |  | Quetiapine | N05AH04 |
| Warfarin | B01AA03 |  |  | Scopolamine | A04AD01, A04AD51,  N05CM05 |
|  |  |  |  | Thioridazine | N05AC02 |
|  |  |  |  | Tolterodine | G04BD07 |
|  |  |  |  | Trifluoperazine | N05AB06 |
|  |  |  |  | Trihexyphenidy | N04AA01 |
|  |  |  |  | Trimipramine | N06AA06 |

^1^Boustani M, Campbell N, Munger S, Maidment I, Fox C. Impact of anticholinergics on the aging brain: a review and practical application. *Aging Health.* 2008/06/01 2008;4(3):311-320. doi:10.2217/1745509X.4.3.311

**Supplementary table 4. Overview of outcomes**

| **Outcomes** | **Explanation** |
| --- | --- |
| **(i) Volume of dispensing outcomes** |  |
| Dispensings | Rate of prescription dispensings per 1,000 population by age and sex |
| Standard daily doses | Rate of dosage units, multiplied by the strength in standard units (i.e. Oral Morphine Equivalents and WHO Defined Daily Doses) per 1,000 population by age and sex |
| Cost | Rate of costs to the Health Service Executive per 1,000 population by age and sex |
| **(ii) Pattern of dispensing outcomes** |  |
| Prevalence of use | Proportion of individuals dispensed a relevant medicine |
| Prevalence of initiations | Proportion of individuals dispensed a relevant medicine with no use in the previous 90 or 180 days |
| Prevalence of discontinuations | Proportion of individuals dispensed a relevant medicine with no further dispensings in the following 90 or 180 days |
| Chronic use | Proportion of individuals dispensed a relevant medicine for >30 days and for >90 days, calculated as a rolling sum over the last 30/90 days, where two or more dispensings in 30 days and four or more dispensings in 90 days equate chronic use |
| **(iii) High-risk dispensings^a^** |  |
| NSAIDs + medications increasing the risk of bleeding | NSAIDs with any of the following: antiplatelet, oral anticoagulant, oral corticosteroids, or SSRIs^b^ |
| NSAIDs + medications increasing the risk of acute kidney injury | NSAIDs with ACEi/ARB and diuretics |
| NSAIDs + medications increasing the risk of acute kidney injury | NSAIDs with metformin and ACEi/ARB in people aged ≥65 years |
| NSAIDs + medication increasing the risk of seizures and neurotoxicity | NSAIDs with lithium |
| Opioids + medication increasing the risk of falls, fractures, and delirium | Opioids with two or more other sedating or anticholinergic^c^ drugs in people aged ≥65 years, |
| Opioids at a dose increasing the risk of dependency | At dose equivalent to >90 mg morphine per day^d^ |
| Gabapentinoids at a dose increasing the risk of dependency | At dose of >4800 mg gabapentin or >800 mg pregabalin per day over last 6 months |

^a^based on Scottish Polypharmacy Guidance^1^

^b^SSRIs and NSAID combination not included in Scottish Polypharmacy guidance, however evidence suggests an elevated bleeding risk^2^

^c^ Anticholinergic drugs identified using the Anticholinergic Cognitive Burden (ACB) scale^3^

1.	Polypharmacy Guidance, Realistic Prescribing (Scottish Government) (2018).

2.	Anglin R, Yuan Y, Moayyedi P, Tse F, Armstrong D, Leontiadis GI. Risk of upper gastrointestinal bleeding with selective serotonin reuptake inhibitors with or without concurrent nonsteroidal anti-inflammatory use: a systematic review and meta-analysis. *Am J Gastroenterol*. Jun 2014;109(6):811-9. doi:10.1038/ajg.2014.82

3.	Boustani M, Campbell N, Munger S, Maidment I, Fox C. Impact of anticholinergics on the aging brain: a review and practical application. *Aging Health*. 2008/06/01 2008;4(3):311-320. doi:10.2217/1745509X.4.3.311

**Supplementary table 5. GMS population 2014-2022**

|  | **GMS** |
| --- | --- |
| **2014** | 1,768,700 |
| **2015** | 1,734,853 |
| **2016** | 1,683,792 |
| **2017** | 1,609,820 |
| **2018** | 1,565,049 |
| **2019** | 1,544,374 |
| **2020** | 1,584,790 |
| **2021** | 1,545,222 |
| **2022** | 1,568,379 |

**Supplementary table 6. Prevalence of any dispensings, initiations (90/180 days), discontinuations (90/180 days), and chronic use (30/90 days) in the GMS population**

**Prevalence of any dispensings**

|  | **2014** | **2015** | **2016** | **2017** | **2018** | **2019** | **2020** | **2021** | **2022** |
| --- | --- | --- | --- | --- | --- | --- | --- | --- | --- |
| **Opioids** | 0.197 | 0.196 | 0.203 | 0.205 | 0.209 | 0.210 | 0.197 | 0.208 | 0.208 |
| *Strong opioids^1^* | 0.093 | 0.095 | 0.098 | 0.099 | 0.099 | 0.099 | 0.093 | 0.098 | 0.094 |
| *Long-acting opioids^2^* | 0.030 | 0.031 | 0.035 | 0.037 | 0.038 | 0.039 | 0.038 | 0.040 | 0.040 |
| *Morphine* | 0.007 | 0.007 | 0.008 | 0.008 | 0.009 | 0.009 | 0.010 | 0.010 | 0.010 |
| *Hydromorphone* | <0.001 | <0.001 | <0.001 | <0.001 | <0.001 | <0.001 | <0.001 | <0.001 | <0.001 |
| *Oxycodone* | 0.012 | 0.013 | 0.015 | 0.017 | 0.018 | 0.019 | 0.018 | 0.021 | 0.021 |
| *Dihydrocodeine* | 0.011 | 0.004 | <0.001 | <0.001 | <0.001 | <0.001 | <0.001 | <0.001 | <0.001 |
| *Pethidine* | <0.001 | <0.001 | <0.001 | <0.001 | <0.001 | <0.001 | 0 | 0 | 0 |
| *Fentanyl* | 0.005 | 0.005 | 0.005 | 0.005 | 0.005 | 0.005 | 0.004 | 0.005 | 0.004 |
| *Buprenorphine* | 0.011 | 0.011 | 0.012 | 0.013 | 0.014 | 0.014 | 0.013 | 0.014 | 0.014 |
| *Tramadol* | 0.073 | 0.072 | 0.071 | 0.068 | 0.065 | 0.062 | 0.055 | 0.056 | 0.054 |
| *Meptazinol* | 0.001 | 0.001 | 0.001 | 0.001 | 0.001 | 0.001 | 0.001 | 0.001 | 0.001 |
| *Tapentadol* | 0.003 | 0.005 | 0.008 | 0.010 | 0.011 | 0.012 | 0.013 | 0.014 | 0.014 |
| *Codeine* | 0.128 | 0.133 | 0.139 | 0.140 | 0.144 | 0.147 | 0.137 | 0.146 | 0.149 |
| **Antimigraine** | 0.011 | 0.011 | 0.011 | 0.011 | 0.011 | 0.010 | 0.010 | 0.010 | 0.010 |
| **Topical analgesics** | 0.140 | 0.147 | 0.158 | 0.164 | 0.151 | 0.152 | 0.135 | 0.147 | 0.149 |
| *Topical NSAID* | 0.126 | 0.129 | 0.135 | 0.143 | 0.148 | 0.149 | 0.133 | 0.145 | 0.147 |
| *Lidocaine* | 0.023 | 0.030 | 0.040 | 0.037 | 0.002 | 0.002 | 0.002 | 0.002 | 0.002 |
| *Capsaicin* | 0.003 | 0.003 | 0.003 | 0.004 | 0.005 | 0.003 | 0.002 | 0.003 | 0.003 |
| **Pregabalin** | 0.044 | 0.046 | 0.049 | 0.050 | 0.048 | 0.046 | 0.043 | 0.045 | 0.045 |
| **Gabapentin** | 0.008 | 0.009 | 0.010 | 0.011 | 0.012 | 0.013 | 0.013 | 0.015 | 0.016 |
| **Amitriptyline** | 0.024 | 0.025 | 0.027 | 0.029 | 0.031 | 0.032 | 0.032 | 0.035 | 0.035 |
| **Paracetamol (inc. combinations)** | 0.297 | 0.296 | 0.307 | 0.310 | 0.321 | 0.325 | 0.299 | 0.311 | 0.320 |
| *Paracetamol (exc. opioid combinations)* | 0.197 | 0.199 | 0.206 | 0.210 | 0.220 | 0.224 | 0.206 | 0.211 | 0.219 |
| **Systemic NSAIDs** | 0.294 | 0.289 | 0.291 | 0.282 | 0.278 | 0.268 | 0.233 | 0.247 | 0.250 |
| *Non-selective NSAIDs* | 0.273 | 0.268 | 0.269 | 0.260 | 0.257 | 0.247 | 0.214 | 0.227 | 0.231 |
| *Coxib* | 0.037 | 0.038 | 0.038 | 0.039 | 0.038 | 0.037 | 0.033 | 0.035 | 0.034 |
| **All analgesics** | 0.483 | 0.476 | 0.486 | 0.486 | 0.488 | 0.480 | 0.434 | 0.455 | 0.463 |

^1^Morphine, hydromorphone, oxycodone, pethidine, fentanyl, buprenorphine, tapentadol, and tramadol

^2^ Slow-release oral morphine, hydromorphone, oxycodone, tramadol, and tapentadol and transdermal fentanyl and buprenorphine

**Prevalence of initiations after 90 and 180 days in the GMS population**

|  | **2014** | **2015** | **2016** | **2017** | **2018** | **2019** | **2020** | **2021** | **2022** |
| --- | --- | --- | --- | --- | --- | --- | --- | --- | --- |
| **90 days** |  |  |  |  |  |  |  |  |  |
| **Opioids** | 0.083 | 0.127 | 0.137 | 0.140 | 0.142 | 0.143 | 0.132 | 0.139 | 0.140 |
| *Strong opioids^1^* | 0.042 | 0.063 | 0.066 | 0.067 | 0.066 | 0.065 | 0.061 | 0.064 | 0.061 |
| *Long-acting opioids^2^* | 0.012 | 0.019 | 0.021 | 0.022 | 0.022 | 0.022 | 0.022 | 0.023 | 0.023 |
| *Morphine* | 0.004 | 0.006 | 0.006 | 0.007 | 0.007 | 0.008 | 0.009 | 0.008 | 0.008 |
| *Hydromorphone* | <0.001 | <0.001 | <0.001 | <0.001 | <0.001 | <0.001 | <0.001 | <0.001 | <0.001 |
| *Oxycodone* | 0.006 | 0.009 | 0.011 | 0.012 | 0.013 | 0.014 | 0.013 | 0.016 | 0.015 |
| *Dihydrocodeine* | 0.005 | 0.002 | <0.001 | <0.001 | <0.001 | <0.001 | <0.001 | <0.001 | <0.001 |
| *Pethidine* | <0.001 | <0.001 | <0.001 | <0.001 | <0.001 | <0.001 | 0.000 | 0.000 | 0.000 |
| *Fentanyl* | 0.002 | 0.002 | 0.002 | 0.002 | 0.002 | 0.002 | 0.002 | 0.002 | 0.002 |
| *Buprenorphine* | 0.005 | 0.007 | 0.007 | 0.008 | 0.008 | 0.008 | 0.007 | 0.008 | 0.008 |
| *Tramadol* | 0.034 | 0.051 | 0.052 | 0.049 | 0.047 | 0.044 | 0.039 | 0.039 | 0.037 |
| *Meptazinol* | <0.001 | <0.001 | 0.001 | 0.001 | 0.001 | 0.001 | 0.001 | 0.001 | <0.001 |
| *Tapentadol* | 0.002 | 0.004 | 0.006 | 0.008 | 0.009 | 0.009 | 0.010 | 0.010 | 0.010 |
| *Codeine* | 0.057 | 0.094 | 0.102 | 0.104 | 0.108 | 0.110 | 0.101 | 0.108 | 0.110 |
| **Antimigraine** | 0.004 | 0.007 | 0.007 | 0.007 | 0.007 | 0.007 | 0.006 | 0.007 | 0.007 |
| **Topical analgesics** | 0.071 | 0.109 | 0.121 | 0.126 | 0.119 | 0.120 | 0.104 | 0.111 | 0.113 |
| *Topical NSAID* | 0.063 | 0.097 | 0.105 | 0.114 | 0.117 | 0.118 | 0.103 | 0.110 | 0.112 |
| *Lidocaine* | 0.013 | 0.024 | 0.031 | 0.027 | 0.002 | 0.001 | 0.001 | 0.001 | 0.001 |
| *Capsaicin* | 0.001 | 0.002 | 0.002 | 0.003 | 0.004 | 0.003 | 0.002 | 0.002 | 0.003 |
| **Pregabalin** | 0.016 | 0.024 | 0.025 | 0.024 | 0.022 | 0.021 | 0.018 | 0.019 | 0.019 |
| **Gabapentin** | 0.003 | 0.005 | 0.005 | 0.006 | 0.007 | 0.007 | 0.007 | 0.008 | 0.009 |
| **Amitriptyline** | 0.008 | 0.013 | 0.013 | 0.014 | 0.015 | 0.016 | 0.015 | 0.016 | 0.016 |
| **Paracetamol (inc. combinations)** | 0.122 | 0.186 | 0.203 | 0.209 | 0.216 | 0.217 | 0.195 | 0.199 | 0.204 |
| *Paracetamol (exc. opioid combinations)* | 0.086 | 0.132 | 0.143 | 0.148 | 0.155 | 0.157 | 0.140 | 0.140 | 0.145 |
| **Systemic NSAIDs** | 0.123 | 0.195 | 0.214 | 0.216 | 0.216 | 0.212 | 0.186 | 0.197 | 0.198 |
| *Non-selective NSAIDs* | 0.114 | 0.182 | 0.200 | 0.201 | 0.202 | 0.199 | 0.174 | 0.184 | 0.186 |
| *Coxib* | 0.019 | 0.028 | 0.030 | 0.031 | 0.030 | 0.030 | 0.026 | 0.027 | 0.027 |
| **All analgesics** | 0.164 | 0.259 | 0.286 | 0.292 | 0.294 | 0.290 | 0.255 | 0.263 | 0.266 |
| **180 days** |  |  |  |  |  |  |  |  |  |
| **Opioids** | 0.040 | 0.101 | 0.113 | 0.116 | 0.118 | 0.119 | 0.110 | 0.116 | 0.117 |
| *Strong opioids^1^* | 0.022 | 0.053 | 0.057 | 0.058 | 0.057 | 0.056 | 0.053 | 0.055 | 0.053 |
| *Long-acting opioids^2^* | 0.007 | 0.017 | 0.018 | 0.020 | 0.020 | 0.020 | 0.019 | 0.021 | 0.021 |
| *Morphine* | 0.002 | 0.005 | 0.006 | 0.006 | 0.007 | 0.007 | 0.008 | 0.008 | 0.008 |
| *Hydromorphone* | <0.001 | <0.001 | <0.001 | <0.001 | <0.001 | <0.001 | <0.001 | <0.001 | <0.001 |
| *Oxycodone* | 0.003 | 0.008 | 0.010 | 0.011 | 0.012 | 0.012 | 0.012 | 0.015 | 0.014 |
| *Dihydrocodeine* | 0.002 | 0.001 | <0.001 | <0.001 | <0.001 | <0.001 | <0.001 | <0.001 | <0.001 |
| *Pethidine* | <0.001 | <0.001 | <0.001 | <0.001 | <0.001 | <0.001 | 0 | 0 | 0 |
| *Fentanyl* | 0.001 | 0.002 | 0.002 | 0.002 | 0.002 | 0.002 | 0.002 | 0.002 | 0.002 |
| *Buprenorphine* | 0.003 | 0.006 | 0.007 | 0.007 | 0.007 | 0.007 | 0.007 | 0.007 | 0.007 |
| *Tramadol* | 0.018 | 0.043 | 0.045 | 0.043 | 0.040 | 0.038 | 0.034 | 0.034 | 0.032 |
| *Meptazinol* | <0.001 | <0.001 | 0.001 | 0.001 | 0.001 | 0.001 | 0.001 | 0.001 | <0.001 |
| *Tapentadol* | 0.001 | 0.004 | 0.006 | 0.007 | 0.008 | 0.009 | 0.009 | 0.010 | 0.009 |
| *Codeine* | 0.029 | 0.077 | 0.086 | 0.088 | 0.092 | 0.094 | 0.087 | 0.093 | 0.095 |
| **Antimigraine** | 0.002 | 0.005 | 0.006 | 0.006 | 0.005 | 0.005 | 0.005 | 0.005 | 0.005 |
| **Topical analgesics** | 0.036 | 0.091 | 0.102 | 0.106 | 0.100 | 0.101 | 0.088 | 0.094 | 0.095 |
| *Topical NSAID* | 0.032 | 0.081 | 0.090 | 0.098 | 0.100 | 0.100 | 0.087 | 0.093 | 0.094 |
| *Lidocaine* | 0.008 | 0.021 | 0.027 | 0.022 | 0.001 | 0.001 | 0.001 | 0.001 | 0.001 |
| *Capsaicin* | 0.001 | 0.002 | 0.002 | 0.003 | 0.004 | 0.003 | 0.001 | 0.002 | 0.003 |
| **Pregabalin** | 0.009 | 0.021 | 0.022 | 0.020 | 0.019 | 0.017 | 0.015 | 0.016 | 0.016 |
| **Gabapentin** | 0.002 | 0.004 | 0.005 | 0.006 | 0.006 | 0.006 | 0.006 | 0.008 | 0.008 |
| **Amitriptyline** | 0.004 | 0.011 | 0.011 | 0.012 | 0.013 | 0.013 | 0.013 | 0.014 | 0.014 |
| **Paracetamol (inc. combinations)** | 0.055 | 0.137 | 0.155 | 0.161 | 0.166 | 0.167 | 0.148 | 0.154 | 0.157 |
| *Paracetamol (exc. opioid combinations)* | 0.041 | 0.101 | 0.112 | 0.117 | 0.122 | 0.124 | 0.108 | 0.110 | 0.113 |
| **Systemic NSAIDs** | 0.057 | 0.151 | 0.174 | 0.177 | 0.179 | 0.176 | 0.155 | 0.165 | 0.166 |
| *Non-selective NSAIDs* | 0.054 | 0.143 | 0.165 | 0.168 | 0.170 | 0.168 | 0.147 | 0.157 | 0.158 |
| *Coxib* | 0.010 | 0.024 | 0.026 | 0.027 | 0.026 | 0.026 | 0.023 | 0.024 | 0.024 |
| **All analgesics** | 0.065 | 0.174 | 0.204 | 0.212 | 0.214 | 0.212 | 0.186 | 0.196 | 0.197 |

^1^Morphine, hydromorphone, oxycodone, pethidine, fentanyl, buprenorphine, tapentadol, and tramadol

^2^ Slow-release oral morphine, hydromorphone, oxycodone, tramadol, and tapentadol and transdermal fentanyl and buprenorphine

**Prevalence of discontinuations after 90 and 180 days in the GMS population**

|  | **2014** | **2015** | **2016** | **2017** | **2018** | **2019** | **2020** | **2021** | **2022** |
| --- | --- | --- | --- | --- | --- | --- | --- | --- | --- |
| **90 days** |  |  |  |  |  |  |  |  |  |
| **Opioids** | 0.070 | 0.105 | 0.113 | 0.116 | 0.117 | 0.118 | 0.107 | 0.113 | 0.115 |
| *Strong opioids^1^* | 0.036 | 0.052 | 0.056 | 0.057 | 0.056 | 0.055 | 0.049 | 0.052 | 0.052 |
| *Long-acting opioids^2^* | 0.009 | 0.014 | 0.016 | 0.017 | 0.017 | 0.018 | 0.016 | 0.018 | 0.018 |
| *Morphine* | 0.002 | 0.002 | 0.002 | 0.003 | 0.003 | 0.003 | 0.003 | 0.003 | 0.003 |
| *Hydromorphone* | <0.001 | <0.001 | <0.001 | <0.001 | <0.001 | <0.001 | <0.001 | <0.001 | <0.001 |
| *Oxycodone* | 0.005 | 0.007 | 0.009 | 0.010 | 0.011 | 0.011 | 0.010 | 0.013 | 0.013 |
| *Dihydrocodeine* | 0.005 | 0.006 | 0.001 | <0.001 | <0.001 | <0.001 | <0.001 | <0.001 | <0.001 |
| *Pethidine* | <0.001 | <0.001 | <0.001 | <0.001 | <0.001 | <0.001 | 0 | 0 | 0 |
| *Fentanyl* | 0.001 | 0.002 | 0.002 | 0.002 | 0.002 | 0.002 | 0.001 | 0.001 | 0.001 |
| *Buprenorphine* | 0.003 | 0.005 | 0.005 | 0.005 | 0.006 | 0.006 | 0.005 | 0.006 | 0.006 |
| *Tramadol* | 0.032 | 0.047 | 0.049 | 0.047 | 0.045 | 0.042 | 0.037 | 0.037 | 0.035 |
| *Meptazinol* | <0.001 | <0.001 | <0.001 | 0.001 | 0.001 | 0.001 | 0.001 | 0.001 | <0.001 |
| *Tapentadol* | 0.001 | 0.003 | 0.005 | 0.006 | 0.007 | 0.008 | 0.008 | 0.010 | 0.009 |
| *Codeine* | 0.051 | 0.080 | 0.088 | 0.091 | 0.093 | 0.095 | 0.087 | 0.093 | 0.096 |
| **Antimigraine** | 0.003 | 0.005 | 0.006 | 0.006 | 0.006 | 0.006 | 0.005 | 0.005 | 0.005 |
| **Topical analgesics** | 0.062 | 0.094 | 0.105 | 0.113 | 0.110 | 0.106 | 0.092 | 0.096 | 0.100 |
| *Topical NSAID* | 0.057 | 0.086 | 0.094 | 0.098 | 0.103 | 0.104 | 0.091 | 0.095 | 0.099 |
| *Lidocaine* | 0.010 | 0.018 | 0.024 | 0.031 | 0.013 | 0.002 | 0.001 | 0.001 | 0.001 |
| *Capsaicin* | 0.001 | 0.002 | 0.002 | 0.003 | 0.004 | 0.004 | 0.002 | 0.002 | 0.002 |
| **Pregabalin** | 0.013 | 0.019 | 0.021 | 0.021 | 0.020 | 0.018 | 0.015 | 0.016 | 0.016 |
| **Gabapentin** | 0.002 | 0.004 | 0.004 | 0.005 | 0.006 | 0.006 | 0.006 | 0.007 | 0.007 |
| **Amitriptyline** | 0.007 | 0.010 | 0.012 | 0.012 | 0.012 | 0.013 | 0.012 | 0.013 | 0.013 |
| **Paracetamol (inc. combinations)** | 0.099 | 0.143 | 0.155 | 0.159 | 0.160 | 0.161 | 0.145 | 0.148 | 0.151 |
| *Paracetamol (exc. opioid combinations)* | 0.072 | 0.106 | 0.115 | 0.118 | 0.120 | 0.122 | 0.110 | 0.109 | 0.112 |
| **Systemic NSAIDs** | 0.102 | 0.146 | 0.159 | 0.162 | 0.162 | 0.164 | 0.145 | 0.153 | 0.155 |
| *Non-selective NSAIDs* | 0.095 | 0.138 | 0.150 | 0.152 | 0.153 | 0.156 | 0.137 | 0.144 | 0.146 |
| *Coxib* | 0.018 | 0.026 | 0.028 | 0.028 | 0.029 | 0.028 | 0.024 | 0.026 | 0.025 |
| **All analgesics** | 0.094 | 0.113 | 0.119 | 0.123 | 0.125 | 0.126 | 0.112 | 0.116 | 0.119 |
| **180 days** |  |  |  |  |  |  |  |  |  |
| **Opioids** | 0.034 | 0.080 | 0.089 | 0.093 | 0.093 | 0.093 | 0.086 | 0.089 | 0.091 |
| *Strong opioids^1^* | 0.019 | 0.043 | 0.047 | 0.048 | 0.047 | 0.047 | 0.042 | 0.043 | 0.043 |
| *Long-acting opioids^2^* | 0.005 | 0.011 | 0.013 | 0.014 | 0.015 | 0.015 | 0.014 | 0.015 | 0.015 |
| *Morphine* | 0.001 | 0.002 | 0.002 | 0.002 | 0.002 | 0.002 | 0.003 | 0.002 | 0.002 |
| *Hydromorphone* | <0.001 | <0.001 | <0.001 | <0.001 | <0.001 | <0.001 | <0.001 | <0.001 | <0.001 |
| *Oxycodone* | 0.003 | 0.006 | 0.007 | 0.008 | 0.009 | 0.010 | 0.009 | 0.011 | 0.012 |
| *Dihydrocodeine* | 0.003 | 0.006 | 0.001 | <0.001 | <0.001 | <0.001 | <0.001 | <0.001 | <0.001 |
| *Pethidine* | <0.001 | <0.001 | <0.001 | <0.001 | <0.001 | <0.001 | 0 | 0 | 0 |
| *Fentanyl* | 0.001 | 0.001 | 0.001 | 0.001 | 0.001 | 0.001 | 0.001 | 0.001 | 0.001 |
| *Buprenorphine* | 0.002 | 0.004 | 0.005 | 0.005 | 0.005 | 0.005 | 0.004 | 0.005 | 0.005 |
| *Tramadol* | 0.017 | 0.039 | 0.042 | 0.041 | 0.039 | 0.037 | 0.032 | 0.032 | 0.030 |
| *Meptazinol* | <0.001 | <0.001 | <0.001 | 0.001 | 0.001 | 0.001 | 0.001 | 0.001 | <0.001 |
| *Tapentadol* | 0.001 | 0.002 | 0.004 | 0.006 | 0.006 | 0.007 | 0.007 | 0.008 | 0.008 |
| *Codeine* | 0.026 | 0.063 | 0.072 | 0.076 | 0.076 | 0.078 | 0.073 | 0.076 | 0.078 |
| **Antimigraine** | 0.002 | 0.004 | 0.005 | 0.005 | 0.005 | 0.005 | 0.004 | 0.004 | 0.004 |
| **Topical analgesics** | 0.031 | 0.073 | 0.083 | 0.089 | 0.094 | 0.087 | 0.076 | 0.076 | 0.080 |
| *Topical NSAID* | 0.029 | 0.068 | 0.077 | 0.080 | 0.083 | 0.085 | 0.075 | 0.076 | 0.079 |
| *Lidocaine* | 0.005 | 0.014 | 0.019 | 0.025 | 0.021 | 0.002 | 0.001 | 0.001 | 0.001 |
| *Capsaicin* | 0.001 | 0.002 | 0.002 | 0.002 | 0.003 | 0.003 | 0.002 | 0.002 | 0.002 |
| **Pregabalin** | 0.007 | 0.016 | 0.018 | 0.018 | 0.017 | 0.016 | 0.013 | 0.013 | 0.013 |
| **Gabapentin** | 0.001 | 0.003 | 0.004 | 0.004 | 0.005 | 0.005 | 0.005 | 0.006 | 0.006 |
| **Amitriptyline** | 0.004 | 0.009 | 0.010 | 0.010 | 0.010 | 0.011 | 0.010 | 0.011 | 0.011 |
| **Paracetamol (inc. combinations)** | 0.044 | 0.099 | 0.110 | 0.114 | 0.114 | 0.114 | 0.104 | 0.105 | 0.105 |
| *Paracetamol (exc. opioid combinations)* | 0.035 | 0.077 | 0.086 | 0.088 | 0.090 | 0.090 | 0.082 | 0.081 | 0.081 |
| **Systemic NSAIDs** | 0.049 | 0.109 | 0.123 | 0.127 | 0.128 | 0.130 | 0.118 | 0.120 | 0.123 |
| *Non-selective NSAIDs* | 0.047 | 0.105 | 0.119 | 0.122 | 0.122 | 0.125 | 0.113 | 0.115 | 0.118 |
| *Coxib* | 0.009 | 0.022 | 0.024 | 0.025 | 0.025 | 0.024 | 0.021 | 0.022 | 0.022 |
| **All analgesics** | 0.039 | 0.064 | 0.069 | 0.072 | 0.074 | 0.075 | 0.068 | 0.070 | 0.071 |

^1^Morphine, hydromorphone, oxycodone, pethidine, fentanyl, buprenorphine, tapentadol, and tramadol

^2^ Slow-release oral morphine, hydromorphone, oxycodone, tramadol, and tapentadol and transdermal fentanyl and buprenorphine

**Prevalence of chronic use (30 and 90 days) in the GMS population**

|  | **2014** | **2015** | **2016** | **2017** | **2018** | **2019** | **2020** | **2021** | **2022** |
| --- | --- | --- | --- | --- | --- | --- | --- | --- | --- |
| **30 days** |  |  |  |  |  |  |  |  |  |
| **Opioids** | 0.089 | 0.092 | 0.096 | 0.100 | 0.103 | 0.105 | 0.105 | 0.111 | 0.109 |
| *Strong opioids^1^* | 0.046 | 0.049 | 0.052 | 0.054 | 0.056 | 0.056 | 0.055 | 0.058 | 0.056 |
| *Long-acting opioids^2^* | 0.020 | 0.021 | 0.024 | 0.025 | 0.026 | 0.027 | 0.026 | 0.027 | 0.026 |
| *Morphine* | 0.004 | 0.004 | 0.004 | 0.004 | 0.005 | 0.005 | 0.005 | 0.005 | 0.005 |
| *Hydromorphone* | <0.001 | <0.001 | <0.001 | <0.001 | <0.001 | <0.001 | <0.001 | <0.001 | <0.001 |
| *Oxycodone* | 0.007 | 0.008 | 0.009 | 0.010 | 0.011 | 0.011 | 0.011 | 0.012 | 0.012 |
| *Dihydrocodeine* | 0.004 | 0.002 | <0.001 | <0.001 | <0.001 | <0.001 | <0.001 | <0.001 | 0 |
| *Pethidine* | <0.001 | <0.001 | <0.001 | <0.001 | <0.001 | <0.001 | 0 | 0 | 0 |
| *Fentanyl* | 0.003 | 0.003 | 0.004 | 0.004 | 0.004 | 0.004 | 0.003 | 0.003 | 0.003 |
| *Buprenorphine* | 0.007 | 0.008 | 0.009 | 0.009 | 0.010 | 0.010 | 0.010 | 0.010 | 0.010 |
| *Tramadol* | 0.029 | 0.029 | 0.030 | 0.029 | 0.028 | 0.027 | 0.026 | 0.026 | 0.025 |
| *Meptazinol* | <0.001 | <0.001 | <0.001 | <0.001 | <0.001 | <0.001 | <0.001 | <0.001 | <0.001 |
| *Tapentadol* | 0.002 | 0.003 | 0.004 | 0.005 | 0.006 | 0.007 | 0.007 | 0.008 | 0.008 |
| *Codeine* | 0.042 | 0.046 | 0.049 | 0.050 | 0.053 | 0.055 | 0.056 | 0.060 | 0.060 |
| **Antimigraine** | 0.004 | 0.005 | 0.005 | 0.005 | 0.005 | 0.005 | 0.005 | 0.005 | 0.005 |
| **Topical analgesics** | 0.043 | 0.049 | 0.055 | 0.058 | 0.047 | 0.050 | 0.052 | 0.058 | 0.058 |
| *Topical NSAID* | 0.033 | 0.036 | 0.038 | 0.042 | 0.045 | 0.048 | 0.050 | 0.056 | 0.057 |
| *Lidocaine* | 0.010 | 0.014 | 0.018 | 0.018 | 0.001 | 0.001 | 0.001 | 0.001 | 0.001 |
| *Capsaicin* | 0.001 | 0.001 | 0.001 | 0.001 | 0.001 | 0.001 | 0.001 | 0.001 | 0.001 |
| **Pregabalin** | 0.030 | 0.033 | 0.035 | 0.036 | 0.036 | 0.035 | 0.033 | 0.034 | 0.034 |
| **Gabapentin** | 0.005 | 0.006 | 0.006 | 0.007 | 0.008 | 0.009 | 0.009 | 0.010 | 0.011 |
| **Amitriptyline** | 0.022 | 0.023 | 0.024 | 0.026 | 0.027 | 0.029 | 0.029 | 0.031 | 0.032 |
| **Paracetamol (inc. combinations)** | 0.126 | 0.131 | 0.138 | 0.144 | 0.152 | 0.158 | 0.161 | 0.170 | 0.172 |
| *Paracetamol (exc. opioid combinations)* | 0.073 | 0.078 | 0.082 | 0.087 | 0.093 | 0.098 | 0.101 | 0.106 | 0.109 |
| **Systemic NSAIDs** | 0.082 | 0.083 | 0.083 | 0.081 | 0.081 | 0.081 | 0.077 | 0.082 | 0.080 |
| *Non-selective NSAIDs* | 0.069 | 0.070 | 0.069 | 0.067 | 0.068 | 0.068 | 0.064 | 0.068 | 0.067 |
| *Coxib* | 0.013 | 0.013 | 0.013 | 0.013 | 0.013 | 0.013 | 0.013 | 0.013 | 0.013 |
| **All analgesics** | 0.293 | 0.297 | 0.309 | 0.315 | 0.320 | 0.324 | 0.308 | 0.326 | 0.327 |
| **90 days** |  |  |  |  |  |  |  |  |  |
| **Opioids** | 0.055 | 0.061 | 0.064 | 0.066 | 0.069 | 0.072 | 0.073 | 0.077 | 0.075 |
| *Strong opioids^1^* | 0.030 | 0.033 | 0.036 | 0.038 | 0.039 | 0.040 | 0.039 | 0.041 | 0.039 |
| *Long-acting opioids^2^* | 0.014 | 0.016 | 0.018 | 0.019 | 0.020 | 0.020 | 0.020 | 0.020 | 0.020 |
| *Morphine* | 0.002 | 0.002 | 0.002 | 0.002 | 0.002 | 0.003 | 0.003 | 0.003 | 0.003 |
| *Hydromorphone* | <0.001 | <0.001 | <0.001 | <0.001 | <0.001 | <0.001 | <0.001 | <0.001 | <0.001 |
| *Oxycodone* | 0.005 | 0.005 | 0.006 | 0.006 | 0.007 | 0.007 | 0.007 | 0.007 | 0.007 |
| *Dihydrocodeine* | 0.002 | 0.001 | <0.001 | <0.001 | <0.001 | <0.001 | <0.001 | <0.001 | 0 |
| *Pethidine* | <0.001 | <0.001 | <0.001 | <0.001 | <0.001 | <0.001 | 0 | 0 | 0 |
| *Fentanyl* | 0.002 | 0.003 | 0.003 | 0.003 | 0.003 | 0.003 | 0.003 | 0.003 | 0.003 |
| *Buprenorphine* | 0.005 | 0.006 | 0.007 | 0.007 | 0.008 | 0.008 | 0.008 | 0.008 | 0.008 |
| *Tramadol* | 0.016 | 0.018 | 0.018 | 0.018 | 0.018 | 0.018 | 0.017 | 0.018 | 0.017 |
| *Meptazinol* | <0.001 | <0.001 | <0.001 | <0.001 | <0.001 | <0.001 | <0.001 | <0.001 | <0.001 |
| *Tapentadol* | 0.001 | 0.002 | 0.002 | 0.003 | 0.004 | 0.004 | 0.004 | 0.005 | 0.004 |
| *Codeine* | 0.023 | 0.027 | 0.029 | 0.030 | 0.032 | 0.033 | 0.035 | 0.038 | 0.038 |
| **Antimigraine** | 0.003 | 0.003 | 0.003 | 0.003 | 0.004 | 0.003 | 0.003 | 0.003 | 0.004 |
| **Topical analgesics** | 0.023 | 0.029 | 0.033 | 0.035 | 0.028 | 0.030 | 0.033 | 0.038 | 0.038 |
| *Topical NSAID* | 0.017 | 0.021 | 0.022 | 0.024 | 0.027 | 0.029 | 0.032 | 0.036 | 0.037 |
| *Lidocaine* | 0.005 | 0.008 | 0.010 | 0.012 | 0.001 | 0.001 | 0.001 | 0.001 | 0.001 |
| *Capsaicin* | <0.001 | <0.001 | <0.001 | <0.001 | 0.001 | 0.001 | <0.001 | <0.001 | <0.001 |
| **Pregabalin** | 0.022 | 0.025 | 0.028 | 0.029 | 0.029 | 0.029 | 0.028 | 0.029 | 0.028 |
| **Gabapentin** | 0.004 | 0.004 | 0.005 | 0.005 | 0.006 | 0.006 | 0.007 | 0.008 | 0.008 |
| **Amitriptyline** | 0.016 | 0.017 | 0.019 | 0.020 | 0.021 | 0.023 | 0.023 | 0.025 | 0.025 |
| **Paracetamol (inc. combinations)** | 0.075 | 0.085 | 0.090 | 0.094 | 0.101 | 0.106 | 0.112 | 0.121 | 0.122 |
| *Paracetamol (exc. opioid combinations)* | 0.042 | 0.050 | 0.053 | 0.056 | 0.060 | 0.064 | 0.070 | 0.076 | 0.077 |
| **Systemic NSAIDs** | 0.032 | 0.036 | 0.035 | 0.035 | 0.035 | 0.035 | 0.036 | 0.038 | 0.036 |
| *Non-selective NSAIDs* | 0.025 | 0.028 | 0.027 | 0.027 | 0.027 | 0.027 | 0.027 | 0.029 | 0.028 |
| *Coxib* | 0.006 | 0.007 | 0.007 | 0.007 | 0.007 | 0.007 | 0.007 | 0.008 | 0.007 |
| **All analgesics** | 0.178 | 0.193 | 0.202 | 0.210 | 0.215 | 0.220 | 0.219 | 0.233 | 0.233 |

^1^Morphine, hydromorphone, oxycodone, pethidine, fentanyl, buprenorphine, tapentadol, and tramadol

^2^ Slow-release oral morphine, hydromorphone, oxycodone, tramadol, and tapentadol and transdermal fentanyl and buprenorphine

**Supplementary table 7. Prevalence of any dispensings by age and sex**

**Prevalence of any dispensings by age group**

|  | **<5** | **5-11** | **12-15** | **16-24** | **25-34** | **35-44** | **45-54** | **55-64** | **65-69** | **70-74** | **75+** |
| --- | --- | --- | --- | --- | --- | --- | --- | --- | --- | --- | --- |
| **2014** |  |  |  |  |  |  |  |  |  |  |  |
| Opioids | 0.002 | 0.003 | 0.023 | 0.123 | 0.200 | 0.228 | 0.261 | 0.298 | 0.297 | 0.297 | 0.352 |
| Strong opioids^1^ | 0.001 | 0.001 | 0.003 | 0.034 | 0.073 | 0.093 | 0.122 | 0.152 | 0.156 | 0.156 | 0.206 |
| Long-acting opioids^2^ | 0.000 | 0.000 | 0.000 | 0.003 | 0.011 | 0.019 | 0.031 | 0.047 | 0.052 | 0.056 | 0.096 |
| Morphine | 0.001 | 0.000 | 0.000 | 0.000 | 0.001 | 0.002 | 0.004 | 0.009 | 0.011 | 0.012 | 0.029 |
| Hydromorphone | 0.000 | 0.000 | 0.000 | 0.000 | 0.000 | 0.000 | 0.001 | 0.001 | 0.001 | 0.001 | 0.001 |
| Oxycodone | 0.000 | 0.000 | 0.000 | 0.003 | 0.007 | 0.010 | 0.016 | 0.022 | 0.024 | 0.023 | 0.026 |
| Dihydrocodeine | 0.000 | 0.000 | 0.001 | 0.004 | 0.007 | 0.009 | 0.013 | 0.018 | 0.020 | 0.019 | 0.024 |
| Pethidine | 0.000 | 0.000 | 0.000 | 0.000 | 0.000 | 0.000 | 0.000 | 0.000 | 0.000 | 0.000 | 0.000 |
| Fentanyl | 0.000 | 0.000 | 0.000 | 0.000 | 0.001 | 0.002 | 0.004 | 0.007 | 0.009 | 0.009 | 0.018 |
| Buprenorphine | 0.000 | 0.000 | 0.000 | 0.000 | 0.001 | 0.003 | 0.005 | 0.011 | 0.014 | 0.019 | 0.052 |
| Tramadol | 0.000 | 0.001 | 0.003 | 0.031 | 0.067 | 0.083 | 0.105 | 0.124 | 0.123 | 0.120 | 0.127 |
| Meptazinol | 0.000 | 0.000 | 0.000 | 0.000 | 0.000 | 0.000 | 0.001 | 0.001 | 0.001 | 0.001 | 0.002 |
| Tapentadol | 0.000 | 0.000 | 0.000 | 0.000 | 0.002 | 0.003 | 0.005 | 0.006 | 0.006 | 0.006 | 0.007 |
| Codeine | 0.001 | 0.002 | 0.019 | 0.098 | 0.150 | 0.164 | 0.174 | 0.189 | 0.182 | 0.180 | 0.194 |
| Antimigraine | 0.000 | 0.001 | 0.003 | 0.012 | 0.017 | 0.018 | 0.024 | 0.016 | 0.009 | 0.006 | 0.003 |
| Topical analgesics | 0.001 | 0.008 | 0.037 | 0.060 | 0.097 | 0.129 | 0.167 | 0.211 | 0.230 | 0.250 | 0.323 |
| Topical NSAID | 0.001 | 0.007 | 0.036 | 0.057 | 0.090 | 0.118 | 0.151 | 0.189 | 0.207 | 0.224 | 0.284 |
| Lidocaine | 0.000 | 0.000 | 0.001 | 0.004 | 0.010 | 0.016 | 0.024 | 0.035 | 0.038 | 0.042 | 0.071 |
| Capsaicin | 0.000 | 0.000 | 0.000 | 0.000 | 0.001 | 0.002 | 0.003 | 0.005 | 0.005 | 0.006 | 0.007 |
| Pregabalin | 0.000 | 0.000 | 0.000 | 0.009 | 0.030 | 0.046 | 0.065 | 0.079 | 0.077 | 0.077 | 0.089 |
| Gabapentin | 0.000 | 0.000 | 0.001 | 0.001 | 0.004 | 0.007 | 0.012 | 0.015 | 0.016 | 0.015 | 0.016 |
| Amitriptyline | 0.000 | 0.000 | 0.001 | 0.007 | 0.018 | 0.028 | 0.040 | 0.047 | 0.045 | 0.041 | 0.040 |
| Paracetamol (inc. combinations) | 0.080 | 0.022 | 0.083 | 0.209 | 0.277 | 0.303 | 0.339 | 0.397 | 0.422 | 0.444 | 0.597 |
| Paracetamol (exc. opioid combinations) | 0.080 | 0.021 | 0.067 | 0.127 | 0.151 | 0.166 | 0.192 | 0.245 | 0.281 | 0.311 | 0.475 |
| Systemic NSAIDs | 0.122 | 0.108 | 0.160 | 0.281 | 0.354 | 0.379 | 0.392 | 0.384 | 0.353 | 0.335 | 0.288 |
| Non-selective NSAIDs | 0.122 | 0.108 | 0.160 | 0.276 | 0.343 | 0.362 | 0.365 | 0.345 | 0.311 | 0.291 | 0.244 |
| Coxib | 0.000 | 0.000 | 0.001 | 0.010 | 0.024 | 0.037 | 0.056 | 0.072 | 0.074 | 0.073 | 0.066 |
| All analgesics | 0.175 | 0.126 | 0.219 | 0.389 | 0.488 | 0.528 | 0.577 | 0.625 | 0.635 | 0.653 | 0.771 |
| **2015** |  |  |  |  |  |  |  |  |  |  |  |
| Opioids | 0.002 | 0.003 | 0.021 | 0.124 | 0.195 | 0.219 | 0.257 | 0.296 | 0.299 | 0.290 | 0.345 |
| Strong opioids^1^ | 0.001 | 0.001 | 0.003 | 0.033 | 0.072 | 0.090 | 0.120 | 0.151 | 0.157 | 0.155 | 0.205 |
| Long-acting opioids^2^ | 0.000 | 0.000 | 0.000 | 0.003 | 0.012 | 0.019 | 0.033 | 0.049 | 0.055 | 0.057 | 0.098 |
| Morphine | 0.001 | 0.001 | 0.001 | 0.000 | 0.001 | 0.002 | 0.004 | 0.008 | 0.011 | 0.012 | 0.031 |
| Hydromorphone | 0.000 | 0.000 | 0.000 | 0.000 | 0.000 | 0.000 | 0.001 | 0.001 | 0.001 | 0.001 | 0.001 |
| Oxycodone | 0.000 | 0.000 | 0.000 | 0.003 | 0.008 | 0.011 | 0.017 | 0.024 | 0.026 | 0.025 | 0.028 |
| Dihydrocodeine | 0.000 | 0.000 | 0.000 | 0.001 | 0.002 | 0.003 | 0.004 | 0.006 | 0.008 | 0.007 | 0.009 |
| Pethidine | 0.000 | 0.000 | 0.000 | 0.000 | 0.000 | 0.000 | 0.000 | 0.000 | 0.000 | 0.000 | 0.000 |
| Fentanyl | 0.000 | 0.000 | 0.000 | 0.000 | 0.001 | 0.002 | 0.004 | 0.007 | 0.008 | 0.008 | 0.017 |
| Buprenorphine | 0.000 | 0.000 | 0.000 | 0.000 | 0.001 | 0.003 | 0.006 | 0.011 | 0.015 | 0.019 | 0.054 |
| Tramadol | 0.000 | 0.001 | 0.003 | 0.029 | 0.065 | 0.079 | 0.101 | 0.121 | 0.121 | 0.116 | 0.121 |
| Meptazinol | 0.000 | 0.000 | 0.000 | 0.000 | 0.000 | 0.000 | 0.001 | 0.001 | 0.001 | 0.001 | 0.002 |
| Tapentadol | 0.000 | 0.000 | 0.000 | 0.001 | 0.003 | 0.005 | 0.007 | 0.009 | 0.009 | 0.009 | 0.010 |
| Codeine | 0.001 | 0.002 | 0.018 | 0.102 | 0.149 | 0.162 | 0.180 | 0.198 | 0.195 | 0.187 | 0.201 |
| Antimigraine | 0.000 | 0.001 | 0.003 | 0.013 | 0.017 | 0.019 | 0.023 | 0.016 | 0.009 | 0.005 | 0.003 |
| Topical analgesics | 0.001 | 0.008 | 0.036 | 0.062 | 0.098 | 0.130 | 0.173 | 0.219 | 0.241 | 0.255 | 0.332 |
| Topical NSAID | 0.001 | 0.007 | 0.035 | 0.058 | 0.089 | 0.115 | 0.152 | 0.192 | 0.210 | 0.223 | 0.284 |
| Lidocaine | 0.000 | 0.000 | 0.001 | 0.005 | 0.013 | 0.022 | 0.032 | 0.045 | 0.051 | 0.055 | 0.091 |
| Capsaicin | 0.000 | 0.000 | 0.000 | 0.000 | 0.001 | 0.002 | 0.003 | 0.005 | 0.005 | 0.006 | 0.007 |
| Pregabalin | 0.000 | 0.000 | 0.000 | 0.010 | 0.032 | 0.049 | 0.067 | 0.082 | 0.081 | 0.079 | 0.091 |
| Gabapentin | 0.000 | 0.000 | 0.001 | 0.001 | 0.004 | 0.008 | 0.013 | 0.016 | 0.016 | 0.016 | 0.017 |
| Amitriptyline | 0.000 | 0.000 | 0.001 | 0.008 | 0.019 | 0.029 | 0.041 | 0.048 | 0.046 | 0.041 | 0.038 |
| Paracetamol (inc. combinations) | 0.077 | 0.020 | 0.080 | 0.214 | 0.270 | 0.292 | 0.335 | 0.398 | 0.426 | 0.433 | 0.579 |
| Paracetamol (exc. opioid combinations) | 0.077 | 0.019 | 0.065 | 0.131 | 0.149 | 0.161 | 0.192 | 0.250 | 0.289 | 0.308 | 0.467 |
| Systemic NSAIDs | 0.117 | 0.102 | 0.155 | 0.297 | 0.347 | 0.369 | 0.385 | 0.378 | 0.348 | 0.319 | 0.271 |
| Non-selective NSAIDs | 0.117 | 0.102 | 0.155 | 0.292 | 0.335 | 0.351 | 0.358 | 0.340 | 0.307 | 0.277 | 0.229 |
| Coxib | 0.000 | 0.000 | 0.001 | 0.010 | 0.024 | 0.037 | 0.055 | 0.072 | 0.074 | 0.071 | 0.063 |
| All analgesics | 0.168 | 0.120 | 0.212 | 0.408 | 0.475 | 0.510 | 0.563 | 0.614 | 0.630 | 0.629 | 0.741 |
| **2016** |  |  |  |  |  |  |  |  |  |  |  |
| Opioids | 0.002 | 0.003 | 0.020 | 0.123 | 0.205 | 0.227 | 0.262 | 0.301 | 0.304 | 0.294 | 0.349 |
| Strong opioids^1^ | 0.001 | 0.001 | 0.004 | 0.032 | 0.074 | 0.094 | 0.121 | 0.154 | 0.159 | 0.156 | 0.209 |
| Long-acting opioids^2^ | 0.000 | 0.000 | 0.001 | 0.004 | 0.013 | 0.021 | 0.035 | 0.053 | 0.059 | 0.062 | 0.103 |
| Morphine | 0.001 | 0.001 | 0.001 | 0.000 | 0.001 | 0.002 | 0.004 | 0.008 | 0.011 | 0.012 | 0.033 |
| Hydromorphone | 0.000 | 0.000 | 0.000 | 0.000 | 0.000 | 0.000 | 0.000 | 0.001 | 0.001 | 0.001 | 0.001 |
| Oxycodone | 0.000 | 0.000 | 0.000 | 0.004 | 0.008 | 0.012 | 0.018 | 0.026 | 0.028 | 0.027 | 0.031 |
| Dihydrocodeine | 0.000 | 0.000 | 0.000 | 0.000 | 0.000 | 0.000 | 0.000 | 0.000 | 0.000 | 0.000 | 0.000 |
| Pethidine | 0.000 | 0.000 | 0.000 | 0.000 | 0.000 | 0.000 | 0.000 | 0.000 | 0.000 | 0.000 | 0.000 |
| Fentanyl | 0.000 | 0.000 | 0.000 | 0.000 | 0.001 | 0.002 | 0.004 | 0.007 | 0.008 | 0.008 | 0.016 |
| Buprenorphine | 0.000 | 0.000 | 0.000 | 0.000 | 0.001 | 0.003 | 0.006 | 0.011 | 0.016 | 0.020 | 0.056 |
| Tramadol | 0.000 | 0.000 | 0.003 | 0.028 | 0.066 | 0.080 | 0.098 | 0.118 | 0.118 | 0.112 | 0.117 |
| Meptazinol | 0.000 | 0.000 | 0.000 | 0.000 | 0.000 | 0.000 | 0.001 | 0.001 | 0.001 | 0.001 | 0.002 |
| Tapentadol | 0.000 | 0.000 | 0.000 | 0.001 | 0.005 | 0.007 | 0.011 | 0.014 | 0.015 | 0.014 | 0.016 |
| Codeine | 0.001 | 0.002 | 0.017 | 0.102 | 0.160 | 0.169 | 0.186 | 0.203 | 0.201 | 0.192 | 0.205 |
| Antimigraine | 0.000 | 0.001 | 0.003 | 0.012 | 0.017 | 0.019 | 0.024 | 0.016 | 0.009 | 0.006 | 0.003 |
| Topical analgesics | 0.001 | 0.007 | 0.034 | 0.061 | 0.105 | 0.138 | 0.179 | 0.230 | 0.254 | 0.267 | 0.351 |
| Topical NSAID | 0.001 | 0.007 | 0.034 | 0.056 | 0.092 | 0.119 | 0.153 | 0.195 | 0.218 | 0.228 | 0.292 |
| Lidocaine | 0.000 | 0.000 | 0.001 | 0.006 | 0.019 | 0.028 | 0.041 | 0.057 | 0.062 | 0.067 | 0.114 |
| Capsaicin | 0.000 | 0.000 | 0.000 | 0.000 | 0.001 | 0.002 | 0.003 | 0.005 | 0.006 | 0.006 | 0.007 |
| Pregabalin | 0.000 | 0.000 | 0.001 | 0.010 | 0.035 | 0.051 | 0.069 | 0.085 | 0.083 | 0.081 | 0.094 |
| Gabapentin | 0.000 | 0.000 | 0.001 | 0.002 | 0.005 | 0.008 | 0.013 | 0.018 | 0.018 | 0.018 | 0.019 |
| Amitriptyline | 0.000 | 0.000 | 0.001 | 0.008 | 0.020 | 0.030 | 0.042 | 0.049 | 0.046 | 0.043 | 0.040 |
| Paracetamol (inc. combinations) | 0.075 | 0.021 | 0.079 | 0.215 | 0.290 | 0.303 | 0.341 | 0.403 | 0.433 | 0.442 | 0.589 |
| Paracetamol (exc. opioid combinations) | 0.074 | 0.019 | 0.064 | 0.131 | 0.161 | 0.167 | 0.194 | 0.253 | 0.292 | 0.314 | 0.475 |
| Systemic NSAIDs | 0.114 | 0.101 | 0.153 | 0.292 | 0.364 | 0.375 | 0.387 | 0.379 | 0.348 | 0.315 | 0.264 |
| Non-selective NSAIDs | 0.114 | 0.101 | 0.152 | 0.287 | 0.353 | 0.358 | 0.359 | 0.340 | 0.307 | 0.273 | 0.224 |
| Coxib | 0.000 | 0.000 | 0.001 | 0.010 | 0.025 | 0.037 | 0.055 | 0.071 | 0.072 | 0.070 | 0.061 |
| All analgesics | 0.163 | 0.118 | 0.208 | 0.404 | 0.499 | 0.522 | 0.567 | 0.620 | 0.636 | 0.634 | 0.751 |
| **2017** |  |  |  |  |  |  |  |  |  |  |  |
| Opioids | 0.002 | 0.003 | 0.018 | 0.111 | 0.204 | 0.233 | 0.266 | 0.298 | 0.300 | 0.291 | 0.344 |
| Strong opioids^1^ | 0.002 | 0.002 | 0.004 | 0.029 | 0.075 | 0.095 | 0.122 | 0.151 | 0.156 | 0.153 | 0.205 |
| Long-acting opioids^2^ | 0.000 | 0.000 | 0.001 | 0.004 | 0.014 | 0.023 | 0.036 | 0.055 | 0.060 | 0.062 | 0.104 |
| Morphine | 0.001 | 0.001 | 0.001 | 0.001 | 0.001 | 0.002 | 0.004 | 0.008 | 0.011 | 0.012 | 0.034 |
| Hydromorphone | 0.000 | 0.000 | 0.000 | 0.000 | 0.000 | 0.000 | 0.000 | 0.001 | 0.001 | 0.001 | 0.000 |
| Oxycodone | 0.000 | 0.000 | 0.000 | 0.005 | 0.010 | 0.015 | 0.020 | 0.029 | 0.030 | 0.029 | 0.033 |
| Dihydrocodeine | 0.000 | 0.000 | 0.000 | 0.000 | 0.000 | 0.000 | 0.000 | 0.000 | 0.000 | 0.000 | 0.000 |
| Pethidine | 0.000 | 0.000 | 0.000 | 0.000 | 0.000 | 0.000 | 0.000 | 0.000 | 0.000 | 0.000 | 0.000 |
| Fentanyl | 0.000 | 0.000 | 0.000 | 0.000 | 0.001 | 0.002 | 0.004 | 0.007 | 0.008 | 0.008 | 0.015 |
| Buprenorphine | 0.000 | 0.000 | 0.000 | 0.000 | 0.001 | 0.003 | 0.006 | 0.012 | 0.017 | 0.020 | 0.055 |
| Tramadol | 0.000 | 0.001 | 0.002 | 0.023 | 0.064 | 0.078 | 0.095 | 0.110 | 0.110 | 0.105 | 0.108 |
| Meptazinol | 0.000 | 0.000 | 0.000 | 0.000 | 0.000 | 0.001 | 0.001 | 0.001 | 0.002 | 0.002 | 0.002 |
| Tapentadol | 0.000 | 0.000 | 0.000 | 0.002 | 0.006 | 0.009 | 0.013 | 0.017 | 0.017 | 0.017 | 0.019 |
| Codeine | 0.001 | 0.001 | 0.015 | 0.092 | 0.158 | 0.173 | 0.189 | 0.202 | 0.200 | 0.189 | 0.203 |
| Antimigraine | 0.000 | 0.001 | 0.003 | 0.012 | 0.018 | 0.020 | 0.025 | 0.017 | 0.009 | 0.006 | 0.003 |
| Topical analgesics | 0.001 | 0.007 | 0.033 | 0.058 | 0.107 | 0.143 | 0.186 | 0.232 | 0.257 | 0.269 | 0.353 |
| Topical NSAID | 0.001 | 0.007 | 0.033 | 0.054 | 0.095 | 0.125 | 0.163 | 0.201 | 0.226 | 0.235 | 0.300 |
| Lidocaine | 0.000 | 0.000 | 0.001 | 0.005 | 0.016 | 0.026 | 0.038 | 0.053 | 0.055 | 0.061 | 0.104 |
| Capsaicin | 0.000 | 0.000 | 0.000 | 0.000 | 0.001 | 0.003 | 0.004 | 0.006 | 0.007 | 0.007 | 0.008 |
| Pregabalin | 0.000 | 0.000 | 0.000 | 0.008 | 0.035 | 0.053 | 0.069 | 0.082 | 0.081 | 0.078 | 0.092 |
| Gabapentin | 0.001 | 0.000 | 0.001 | 0.002 | 0.006 | 0.010 | 0.015 | 0.019 | 0.019 | 0.020 | 0.020 |
| Amitriptyline | 0.000 | 0.000 | 0.001 | 0.008 | 0.023 | 0.034 | 0.046 | 0.051 | 0.048 | 0.044 | 0.041 |
| Paracetamol (inc. combinations) | 0.063 | 0.016 | 0.070 | 0.199 | 0.289 | 0.312 | 0.351 | 0.404 | 0.433 | 0.440 | 0.583 |
| Paracetamol (exc. opioid combinations) | 0.063 | 0.015 | 0.058 | 0.124 | 0.161 | 0.171 | 0.199 | 0.254 | 0.292 | 0.314 | 0.473 |
| Systemic NSAIDs | 0.090 | 0.081 | 0.139 | 0.273 | 0.360 | 0.383 | 0.390 | 0.372 | 0.339 | 0.306 | 0.251 |
| Non-selective NSAIDs | 0.090 | 0.081 | 0.139 | 0.268 | 0.349 | 0.365 | 0.362 | 0.333 | 0.297 | 0.264 | 0.211 |
| Coxib | 0.000 | 0.000 | 0.001 | 0.009 | 0.025 | 0.038 | 0.056 | 0.071 | 0.073 | 0.069 | 0.060 |
| All analgesics | 0.135 | 0.096 | 0.192 | 0.379 | 0.500 | 0.539 | 0.582 | 0.620 | 0.632 | 0.627 | 0.740 |
| **2018** |  |  |  |  |  |  |  |  |  |  |  |
| Opioids | 0.002 | 0.003 | 0.017 | 0.110 | 0.204 | 0.233 | 0.263 | 0.299 | 0.308 | 0.294 | 0.346 |
| Strong opioids^1^ | 0.002 | 0.002 | 0.004 | 0.027 | 0.072 | 0.093 | 0.116 | 0.146 | 0.157 | 0.152 | 0.202 |
| Long-acting opioids^2^ | 0.000 | 0.000 | 0.001 | 0.004 | 0.014 | 0.023 | 0.036 | 0.054 | 0.063 | 0.064 | 0.106 |
| Morphine | 0.002 | 0.001 | 0.002 | 0.001 | 0.001 | 0.002 | 0.004 | 0.008 | 0.012 | 0.013 | 0.035 |
| Hydromorphone | 0.000 | 0.000 | 0.000 | 0.000 | 0.000 | 0.000 | 0.000 | 0.001 | 0.001 | 0.000 | 0.000 |
| Oxycodone | 0.000 | 0.000 | 0.000 | 0.004 | 0.011 | 0.015 | 0.020 | 0.029 | 0.032 | 0.031 | 0.034 |
| Dihydrocodeine | 0.000 | 0.000 | 0.000 | 0.000 | 0.000 | 0.000 | 0.000 | 0.000 | 0.000 | 0.000 | 0.000 |
| Pethidine | 0.000 | 0.000 | 0.000 | 0.000 | 0.000 | 0.000 | 0.000 | 0.000 | 0.000 | 0.000 | 0.000 |
| Fentanyl | 0.000 | 0.000 | 0.000 | 0.000 | 0.001 | 0.002 | 0.004 | 0.007 | 0.008 | 0.008 | 0.015 |
| Buprenorphine | 0.000 | 0.000 | 0.000 | 0.000 | 0.001 | 0.003 | 0.006 | 0.012 | 0.018 | 0.021 | 0.057 |
| Tramadol | 0.000 | 0.000 | 0.002 | 0.020 | 0.059 | 0.075 | 0.088 | 0.104 | 0.105 | 0.099 | 0.100 |
| Meptazinol | 0.000 | 0.000 | 0.000 | 0.000 | 0.000 | 0.000 | 0.001 | 0.001 | 0.001 | 0.002 | 0.002 |
| Tapentadol | 0.000 | 0.000 | 0.000 | 0.002 | 0.007 | 0.011 | 0.014 | 0.018 | 0.020 | 0.019 | 0.021 |
| Codeine | 0.001 | 0.001 | 0.014 | 0.092 | 0.161 | 0.175 | 0.191 | 0.206 | 0.209 | 0.197 | 0.209 |
| Antimigraine | 0.000 | 0.001 | 0.004 | 0.012 | 0.018 | 0.021 | 0.024 | 0.016 | 0.009 | 0.006 | 0.003 |
| Topical analgesics | 0.001 | 0.006 | 0.030 | 0.054 | 0.098 | 0.129 | 0.165 | 0.209 | 0.237 | 0.244 | 0.314 |
| Topical NSAID | 0.001 | 0.006 | 0.030 | 0.054 | 0.097 | 0.127 | 0.162 | 0.204 | 0.232 | 0.238 | 0.307 |
| Lidocaine | 0.000 | 0.000 | 0.000 | 0.000 | 0.000 | 0.001 | 0.002 | 0.003 | 0.003 | 0.004 | 0.006 |
| Capsaicin | 0.000 | 0.000 | 0.000 | 0.001 | 0.002 | 0.003 | 0.005 | 0.007 | 0.009 | 0.008 | 0.010 |
| Pregabalin | 0.000 | 0.000 | 0.000 | 0.008 | 0.034 | 0.050 | 0.064 | 0.078 | 0.079 | 0.074 | 0.088 |
| Gabapentin | 0.000 | 0.000 | 0.001 | 0.002 | 0.006 | 0.011 | 0.016 | 0.020 | 0.021 | 0.021 | 0.022 |
| Amitriptyline | 0.000 | 0.000 | 0.001 | 0.009 | 0.023 | 0.036 | 0.047 | 0.052 | 0.051 | 0.045 | 0.043 |
| Paracetamol (inc. combinations) | 0.063 | 0.017 | 0.070 | 0.204 | 0.298 | 0.318 | 0.350 | 0.410 | 0.449 | 0.449 | 0.593 |
| Paracetamol (exc. opioid combinations) | 0.063 | 0.016 | 0.059 | 0.130 | 0.172 | 0.178 | 0.202 | 0.260 | 0.308 | 0.324 | 0.484 |
| Systemic NSAIDs | 0.069 | 0.068 | 0.132 | 0.278 | 0.365 | 0.383 | 0.380 | 0.368 | 0.342 | 0.303 | 0.246 |
| Non-selective NSAIDs | 0.069 | 0.068 | 0.132 | 0.274 | 0.354 | 0.366 | 0.354 | 0.332 | 0.301 | 0.263 | 0.208 |
| Coxib | 0.000 | 0.000 | 0.001 | 0.009 | 0.025 | 0.038 | 0.054 | 0.067 | 0.071 | 0.066 | 0.056 |
| All analgesics | 0.117 | 0.084 | 0.183 | 0.383 | 0.504 | 0.538 | 0.567 | 0.617 | 0.642 | 0.628 | 0.740 |
| **2019** |  |  |  |  |  |  |  |  |  |  |  |
| Opioids | 0.003 | 0.003 | 0.016 | 0.111 | 0.197 | 0.228 | 0.260 | 0.300 | 0.314 | 0.296 | 0.347 |
| Strong opioids^1^ | 0.002 | 0.002 | 0.003 | 0.027 | 0.067 | 0.090 | 0.113 | 0.144 | 0.155 | 0.148 | 0.199 |
| Long-acting opioids^2^ | 0.000 | 0.000 | 0.000 | 0.004 | 0.012 | 0.023 | 0.035 | 0.054 | 0.063 | 0.063 | 0.104 |
| Morphine | 0.002 | 0.001 | 0.001 | 0.001 | 0.001 | 0.002 | 0.004 | 0.009 | 0.012 | 0.013 | 0.036 |
| Hydromorphone | 0.000 | 0.000 | 0.000 | 0.000 | 0.000 | 0.000 | 0.000 | 0.001 | 0.001 | 0.000 | 0.000 |
| Oxycodone | 0.000 | 0.000 | 0.000 | 0.005 | 0.010 | 0.015 | 0.021 | 0.030 | 0.033 | 0.031 | 0.036 |
| Dihydrocodeine | 0.000 | 0.000 | 0.000 | 0.000 | 0.000 | 0.000 | 0.000 | 0.000 | 0.000 | 0.000 | 0.000 |
| Pethidine | 0.000 | 0.000 | 0.000 | 0.000 | 0.000 | 0.000 | 0.000 | 0.000 | 0.000 | 0.000 | 0.000 |
| Fentanyl | 0.000 | 0.000 | 0.000 | 0.000 | 0.001 | 0.002 | 0.004 | 0.006 | 0.007 | 0.007 | 0.014 |
| Buprenorphine | 0.000 | 0.000 | 0.000 | 0.000 | 0.001 | 0.003 | 0.006 | 0.012 | 0.017 | 0.020 | 0.055 |
| Tramadol | 0.000 | 0.000 | 0.002 | 0.020 | 0.055 | 0.070 | 0.084 | 0.099 | 0.101 | 0.092 | 0.093 |
| Meptazinol | 0.000 | 0.000 | 0.000 | 0.000 | 0.000 | 0.000 | 0.001 | 0.001 | 0.001 | 0.002 | 0.002 |
| Tapentadol | 0.000 | 0.000 | 0.000 | 0.002 | 0.007 | 0.012 | 0.015 | 0.020 | 0.022 | 0.020 | 0.022 |
| Codeine | 0.001 | 0.001 | 0.013 | 0.093 | 0.157 | 0.173 | 0.190 | 0.211 | 0.217 | 0.203 | 0.213 |
| Antimigraine | 0.000 | 0.001 | 0.003 | 0.013 | 0.020 | 0.021 | 0.021 | 0.014 | 0.008 | 0.005 | 0.002 |
| Topical analgesics | 0.001 | 0.006 | 0.027 | 0.054 | 0.093 | 0.125 | 0.163 | 0.208 | 0.239 | 0.244 | 0.314 |
| Topical NSAID | 0.001 | 0.006 | 0.027 | 0.054 | 0.092 | 0.123 | 0.159 | 0.204 | 0.235 | 0.240 | 0.308 |
| Lidocaine | 0.000 | 0.000 | 0.000 | 0.000 | 0.000 | 0.001 | 0.001 | 0.003 | 0.003 | 0.003 | 0.006 |
| Capsaicin | 0.000 | 0.000 | 0.000 | 0.000 | 0.001 | 0.002 | 0.004 | 0.005 | 0.006 | 0.006 | 0.007 |
| Pregabalin | 0.000 | 0.000 | 0.000 | 0.007 | 0.032 | 0.046 | 0.060 | 0.074 | 0.077 | 0.070 | 0.084 |
| Gabapentin | 0.001 | 0.000 | 0.001 | 0.002 | 0.006 | 0.011 | 0.016 | 0.021 | 0.023 | 0.022 | 0.023 |
| Amitriptyline | 0.000 | 0.000 | 0.001 | 0.009 | 0.024 | 0.037 | 0.049 | 0.055 | 0.054 | 0.047 | 0.044 |
| Paracetamol (inc. combinations) | 0.063 | 0.018 | 0.071 | 0.211 | 0.292 | 0.313 | 0.350 | 0.413 | 0.460 | 0.455 | 0.595 |
| Paracetamol (exc. opioid combinations) | 0.062 | 0.017 | 0.060 | 0.135 | 0.169 | 0.177 | 0.204 | 0.261 | 0.315 | 0.328 | 0.487 |
| Systemic NSAIDs | 0.026 | 0.030 | 0.111 | 0.281 | 0.352 | 0.372 | 0.379 | 0.369 | 0.347 | 0.302 | 0.239 |
| Non-selective NSAIDs | 0.026 | 0.030 | 0.110 | 0.276 | 0.341 | 0.356 | 0.355 | 0.333 | 0.307 | 0.263 | 0.205 |
| Coxib | 0.000 | 0.000 | 0.001 | 0.009 | 0.024 | 0.035 | 0.052 | 0.066 | 0.069 | 0.065 | 0.052 |
| All analgesics | 0.083 | 0.049 | 0.163 | 0.389 | 0.487 | 0.522 | 0.561 | 0.614 | 0.652 | 0.629 | 0.735 |
| **2020** |  |  |  |  |  |  |  |  |  |  |  |
| Opioids | 0.002 | 0.002 | 0.010 | 0.079 | 0.172 | 0.208 | 0.246 | 0.287 | 0.301 | 0.281 | 0.339 |
| Strong opioids^1^ | 0.002 | 0.001 | 0.003 | 0.019 | 0.060 | 0.082 | 0.107 | 0.136 | 0.148 | 0.139 | 0.194 |
| Long-acting opioids^2^ | 0.000 | 0.000 | 0.000 | 0.003 | 0.011 | 0.021 | 0.034 | 0.052 | 0.062 | 0.061 | 0.103 |
| Morphine | 0.002 | 0.001 | 0.002 | 0.001 | 0.001 | 0.002 | 0.004 | 0.009 | 0.013 | 0.014 | 0.042 |
| Hydromorphone | 0.000 | 0.000 | 0.000 | 0.000 | 0.000 | 0.000 | 0.000 | 0.001 | 0.000 | 0.000 | 0.000 |
| Oxycodone | 0.000 | 0.000 | 0.000 | 0.004 | 0.009 | 0.014 | 0.021 | 0.030 | 0.033 | 0.030 | 0.036 |
| Dihydrocodeine | 0.000 | 0.000 | 0.000 | 0.000 | 0.000 | 0.000 | 0.000 | 0.000 | 0.000 | 0.000 | 0.000 |
| Pethidine | 0.000 | 0.000 | 0.000 | 0.000 | 0.000 | 0.000 | 0.000 | 0.000 | 0.000 | 0.000 | 0.000 |
| Fentanyl | 0.000 | 0.000 | 0.000 | 0.000 | 0.001 | 0.002 | 0.003 | 0.006 | 0.008 | 0.007 | 0.013 |
| Buprenorphine | 0.000 | 0.000 | 0.000 | 0.000 | 0.001 | 0.002 | 0.006 | 0.011 | 0.016 | 0.020 | 0.055 |
| Tramadol | 0.000 | 0.000 | 0.001 | 0.014 | 0.048 | 0.063 | 0.077 | 0.090 | 0.092 | 0.082 | 0.082 |
| Meptazinol | 0.000 | 0.000 | 0.000 | 0.000 | 0.000 | 0.000 | 0.001 | 0.001 | 0.001 | 0.001 | 0.002 |
| Tapentadol | 0.000 | 0.000 | 0.000 | 0.002 | 0.007 | 0.012 | 0.016 | 0.021 | 0.023 | 0.021 | 0.023 |
| Codeine | 0.001 | 0.001 | 0.008 | 0.066 | 0.136 | 0.159 | 0.180 | 0.202 | 0.208 | 0.193 | 0.207 |
| Antimigraine | 0.000 | 0.000 | 0.003 | 0.011 | 0.018 | 0.021 | 0.019 | 0.012 | 0.008 | 0.005 | 0.002 |
| Topical analgesics | 0.000 | 0.004 | 0.017 | 0.037 | 0.076 | 0.106 | 0.145 | 0.187 | 0.218 | 0.227 | 0.290 |
| Topical NSAID | 0.000 | 0.004 | 0.017 | 0.037 | 0.075 | 0.104 | 0.143 | 0.184 | 0.215 | 0.223 | 0.286 |
| Lidocaine | 0.000 | 0.000 | 0.000 | 0.000 | 0.000 | 0.001 | 0.001 | 0.003 | 0.003 | 0.003 | 0.006 |
| Capsaicin | 0.000 | 0.000 | 0.000 | 0.000 | 0.001 | 0.001 | 0.002 | 0.003 | 0.003 | 0.004 | 0.004 |
| Pregabalin | 0.000 | 0.000 | 0.000 | 0.005 | 0.026 | 0.042 | 0.055 | 0.069 | 0.073 | 0.064 | 0.079 |
| Gabapentin | 0.001 | 0.001 | 0.001 | 0.002 | 0.006 | 0.011 | 0.017 | 0.021 | 0.025 | 0.022 | 0.024 |
| Amitriptyline | 0.000 | 0.000 | 0.001 | 0.008 | 0.022 | 0.035 | 0.048 | 0.054 | 0.054 | 0.048 | 0.044 |
| Paracetamol (inc. combinations) | 0.031 | 0.010 | 0.040 | 0.143 | 0.249 | 0.281 | 0.328 | 0.396 | 0.442 | 0.441 | 0.577 |
| Paracetamol (exc. opioid combinations) | 0.030 | 0.009 | 0.033 | 0.089 | 0.140 | 0.155 | 0.188 | 0.250 | 0.303 | 0.319 | 0.472 |
| Systemic NSAIDs | 0.018 | 0.022 | 0.070 | 0.197 | 0.299 | 0.325 | 0.347 | 0.338 | 0.315 | 0.271 | 0.213 |
| Non-selective NSAIDs | 0.018 | 0.022 | 0.070 | 0.194 | 0.291 | 0.310 | 0.323 | 0.303 | 0.277 | 0.236 | 0.181 |
| Coxib | 0.000 | 0.000 | 0.001 | 0.007 | 0.020 | 0.031 | 0.048 | 0.061 | 0.064 | 0.057 | 0.047 |
| All analgesics | 0.047 | 0.034 | 0.101 | 0.271 | 0.411 | 0.460 | 0.520 | 0.579 | 0.616 | 0.600 | 0.707 |
| **2021** |  |  |  |  |  |  |  |  |  |  |  |
| Opioids | 0.003 | 0.002 | 0.010 | 0.090 | 0.186 | 0.219 | 0.257 | 0.297 | 0.315 | 0.292 | 0.343 |
| Strong opioids^1^ | 0.002 | 0.002 | 0.003 | 0.022 | 0.064 | 0.086 | 0.110 | 0.140 | 0.154 | 0.143 | 0.195 |
| Long-acting opioids^2^ | 0.000 | 0.000 | 0.000 | 0.003 | 0.011 | 0.022 | 0.035 | 0.054 | 0.064 | 0.063 | 0.105 |
| Morphine | 0.002 | 0.001 | 0.002 | 0.001 | 0.001 | 0.002 | 0.004 | 0.008 | 0.012 | 0.013 | 0.039 |
| Hydromorphone | 0.000 | 0.000 | 0.000 | 0.000 | 0.000 | 0.000 | 0.000 | 0.001 | 0.000 | 0.000 | 0.000 |
| Oxycodone | 0.000 | 0.000 | 0.000 | 0.005 | 0.012 | 0.016 | 0.023 | 0.033 | 0.038 | 0.034 | 0.040 |
| Dihydrocodeine | 0.000 | 0.000 | 0.000 | 0.000 | 0.000 | 0.000 | 0.000 | 0.000 | 0.000 | 0.000 | 0.000 |
| Pethidine | 0.000 | 0.000 | 0.000 | 0.000 | 0.000 | 0.000 | 0.000 | 0.000 | 0.000 | 0.000 | 0.000 |
| Fentanyl | 0.000 | 0.000 | 0.000 | 0.000 | 0.001 | 0.002 | 0.003 | 0.006 | 0.008 | 0.007 | 0.013 |
| Buprenorphine | 0.000 | 0.000 | 0.000 | 0.000 | 0.001 | 0.003 | 0.006 | 0.011 | 0.017 | 0.021 | 0.057 |
| Tramadol | 0.000 | 0.000 | 0.001 | 0.016 | 0.050 | 0.064 | 0.077 | 0.090 | 0.093 | 0.082 | 0.080 |
| Meptazinol | 0.000 | 0.000 | 0.000 | 0.000 | 0.000 | 0.000 | 0.001 | 0.001 | 0.001 | 0.001 | 0.002 |
| Tapentadol | 0.000 | 0.000 | 0.000 | 0.002 | 0.008 | 0.013 | 0.017 | 0.022 | 0.024 | 0.022 | 0.025 |
| Codeine | 0.000 | 0.001 | 0.007 | 0.076 | 0.149 | 0.169 | 0.190 | 0.211 | 0.219 | 0.202 | 0.213 |
| Antimigraine | 0.000 | 0.001 | 0.003 | 0.013 | 0.020 | 0.024 | 0.020 | 0.012 | 0.008 | 0.005 | 0.002 |
| Topical analgesics | 0.001 | 0.004 | 0.017 | 0.043 | 0.082 | 0.115 | 0.156 | 0.197 | 0.229 | 0.237 | 0.305 |
| Topical NSAID | 0.001 | 0.004 | 0.017 | 0.043 | 0.082 | 0.114 | 0.153 | 0.194 | 0.225 | 0.233 | 0.300 |
| Lidocaine | 0.000 | 0.000 | 0.000 | 0.000 | 0.000 | 0.001 | 0.001 | 0.002 | 0.003 | 0.003 | 0.005 |
| Capsaicin | 0.000 | 0.000 | 0.000 | 0.000 | 0.001 | 0.002 | 0.003 | 0.005 | 0.005 | 0.005 | 0.006 |
| Pregabalin | 0.000 | 0.000 | 0.000 | 0.006 | 0.029 | 0.042 | 0.058 | 0.071 | 0.076 | 0.067 | 0.080 |
| Gabapentin | 0.001 | 0.001 | 0.001 | 0.003 | 0.007 | 0.013 | 0.019 | 0.024 | 0.027 | 0.025 | 0.027 |
| Amitriptyline | 0.000 | 0.000 | 0.001 | 0.009 | 0.024 | 0.038 | 0.052 | 0.058 | 0.057 | 0.050 | 0.047 |
| Paracetamol (inc. combinations) | 0.029 | 0.008 | 0.034 | 0.154 | 0.264 | 0.293 | 0.340 | 0.405 | 0.455 | 0.450 | 0.578 |
| Paracetamol (exc. opioid combinations) | 0.028 | 0.007 | 0.028 | 0.091 | 0.144 | 0.158 | 0.194 | 0.255 | 0.310 | 0.320 | 0.469 |
| Systemic NSAIDs | 0.026 | 0.023 | 0.073 | 0.227 | 0.323 | 0.347 | 0.363 | 0.352 | 0.323 | 0.282 | 0.215 |
| Non-selective NSAIDs | 0.026 | 0.023 | 0.072 | 0.223 | 0.313 | 0.332 | 0.339 | 0.318 | 0.285 | 0.245 | 0.183 |
| Coxib | 0.000 | 0.000 | 0.001 | 0.008 | 0.022 | 0.034 | 0.050 | 0.062 | 0.064 | 0.059 | 0.048 |
| All analgesics | 0.052 | 0.033 | 0.101 | 0.305 | 0.438 | 0.482 | 0.539 | 0.595 | 0.633 | 0.612 | 0.714 |
| **2022** |  |  |  |  |  |  |  |  |  |  |  |
| Opioids | 0.003 | 0.002 | 0.012 | 0.089 | 0.176 | 0.217 | 0.255 | 0.295 | 0.314 | 0.297 | 0.346 |
| Strong opioids^1^ | 0.002 | 0.002 | 0.003 | 0.019 | 0.055 | 0.080 | 0.105 | 0.135 | 0.149 | 0.142 | 0.190 |
| Long-acting opioids^2^ | 0.000 | 0.000 | 0.000 | 0.003 | 0.011 | 0.021 | 0.035 | 0.053 | 0.064 | 0.063 | 0.103 |
| Morphine | 0.002 | 0.001 | 0.002 | 0.001 | 0.001 | 0.002 | 0.004 | 0.008 | 0.013 | 0.013 | 0.037 |
| Hydromorphone | 0.000 | 0.000 | 0.000 | 0.000 | 0.000 | 0.000 | 0.000 | 0.000 | 0.000 | 0.000 | 0.000 |
| Oxycodone | 0.000 | 0.000 | 0.000 | 0.004 | 0.010 | 0.016 | 0.022 | 0.031 | 0.037 | 0.035 | 0.039 |
| Dihydrocodeine | 0.000 | 0.000 | 0.000 | 0.000 | 0.000 | 0.000 | 0.000 | 0.000 | 0.000 | 0.000 | 0.000 |
| Pethidine | 0.000 | 0.000 | 0.000 | 0.000 | 0.000 | 0.000 | 0.000 | 0.000 | 0.000 | 0.000 | 0.000 |
| Fentanyl | 0.000 | 0.000 | 0.000 | 0.000 | 0.001 | 0.001 | 0.003 | 0.006 | 0.007 | 0.007 | 0.012 |
| Buprenorphine | 0.000 | 0.000 | 0.000 | 0.000 | 0.001 | 0.002 | 0.005 | 0.010 | 0.016 | 0.020 | 0.055 |
| Tramadol | 0.000 | 0.000 | 0.001 | 0.013 | 0.042 | 0.059 | 0.073 | 0.086 | 0.089 | 0.080 | 0.077 |
| Meptazinol | 0.000 | 0.000 | 0.000 | 0.000 | 0.000 | 0.000 | 0.000 | 0.001 | 0.001 | 0.001 | 0.002 |
| Tapentadol | 0.000 | 0.000 | 0.000 | 0.002 | 0.007 | 0.011 | 0.016 | 0.021 | 0.023 | 0.023 | 0.024 |
| Codeine | 0.001 | 0.001 | 0.009 | 0.076 | 0.144 | 0.170 | 0.192 | 0.212 | 0.224 | 0.211 | 0.220 |
| Antimigraine | 0.000 | 0.001 | 0.003 | 0.013 | 0.020 | 0.023 | 0.021 | 0.013 | 0.008 | 0.005 | 0.002 |
| Topical analgesics | 0.000 | 0.004 | 0.018 | 0.041 | 0.077 | 0.112 | 0.154 | 0.198 | 0.231 | 0.245 | 0.315 |
| Topical NSAID | 0.000 | 0.004 | 0.018 | 0.041 | 0.076 | 0.111 | 0.152 | 0.194 | 0.227 | 0.241 | 0.310 |
| Lidocaine | 0.000 | 0.000 | 0.000 | 0.000 | 0.000 | 0.001 | 0.001 | 0.002 | 0.003 | 0.003 | 0.005 |
| Capsaicin | 0.000 | 0.000 | 0.000 | 0.000 | 0.001 | 0.002 | 0.003 | 0.005 | 0.005 | 0.006 | 0.007 |
| Pregabalin | 0.000 | 0.000 | 0.000 | 0.005 | 0.025 | 0.041 | 0.057 | 0.070 | 0.074 | 0.067 | 0.080 |
| Gabapentin | 0.001 | 0.001 | 0.001 | 0.003 | 0.007 | 0.012 | 0.021 | 0.025 | 0.028 | 0.026 | 0.028 |
| Amitriptyline | 0.000 | 0.000 | 0.001 | 0.008 | 0.023 | 0.037 | 0.053 | 0.059 | 0.058 | 0.053 | 0.049 |
| Paracetamol (inc. combinations) | 0.040 | 0.012 | 0.048 | 0.163 | 0.257 | 0.299 | 0.345 | 0.410 | 0.459 | 0.463 | 0.589 |
| Paracetamol (exc. opioid combinations) | 0.039 | 0.011 | 0.040 | 0.100 | 0.143 | 0.164 | 0.197 | 0.259 | 0.314 | 0.333 | 0.480 |
| Systemic NSAIDs | 0.041 | 0.036 | 0.087 | 0.230 | 0.313 | 0.344 | 0.361 | 0.350 | 0.321 | 0.287 | 0.219 |
| Non-selective NSAIDs | 0.041 | 0.036 | 0.087 | 0.226 | 0.304 | 0.329 | 0.337 | 0.317 | 0.284 | 0.251 | 0.187 |
| Coxib | 0.000 | 0.000 | 0.001 | 0.007 | 0.020 | 0.032 | 0.048 | 0.059 | 0.062 | 0.059 | 0.047 |
| All analgesics | 0.074 | 0.048 | 0.122 | 0.314 | 0.426 | 0.483 | 0.540 | 0.594 | 0.630 | 0.622 | 0.722 |

**Prevalence of any dispensings by sex**

|  | **Females** | **Males** |
| --- | --- | --- |
| **2014** |  |  |
| Opioids | 0.227 | 0.162 |
| Strong opioids^1^ | 0.107 | 0.079 |
| Long-acting opioids^2^ | 0.035 | 0.024 |
| Morphine | 0.007 | 0.006 |
| Hydromorphone | 0.000 | 0.000 |
| Oxycodone | 0.013 | 0.011 |
| Dihydrocodeine | 0.012 | 0.009 |
| Pethidine | 0.000 | 0.000 |
| Fentanyl | 0.006 | 0.003 |
| Buprenorphine | 0.014 | 0.006 |
| Tramadol | 0.082 | 0.063 |
| Meptazinol | 0.001 | 0.000 |
| Tapentadol | 0.004 | 0.002 |
| Codeine | 0.152 | 0.102 |
| Antimigraine | 0.016 | 0.004 |
| Topical analgesics | 0.165 | 0.112 |
| Topical NSAID | 0.148 | 0.102 |
| Lidocaine | 0.029 | 0.015 |
| Capsaicin | 0.003 | 0.002 |
| Pregabalin | 0.050 | 0.036 |
| Gabapentin | 0.009 | 0.006 |
| Amitriptyline | 0.031 | 0.017 |
| Paracetamol (inc. combinations) | 0.343 | 0.245 |
| Paracetamol (exc. opioid combinations) | 0.230 | 0.160 |
| Systemic NSAIDs | 0.332 | 0.251 |
| Non-selective NSAIDs | 0.310 | 0.232 |
| Coxib | 0.042 | 0.032 |
| All analgesics | 0.540 | 0.419 |
| **2015** |  |  |
| Opioids | 0.227 | 0.161 |
| Strong opioids^1^ | 0.108 | 0.079 |
| Long-acting opioids^2^ | 0.037 | 0.025 |
| Morphine | 0.008 | 0.007 |
| Hydromorphone | 0.000 | 0.000 |
| Oxycodone | 0.015 | 0.012 |
| Dihydrocodeine | 0.004 | 0.003 |
| Pethidine | 0.000 | 0.000 |
| Fentanyl | 0.006 | 0.003 |
| Buprenorphine | 0.015 | 0.007 |
| Tramadol | 0.081 | 0.061 |
| Meptazinol | 0.001 | 0.000 |
| Tapentadol | 0.006 | 0.004 |
| Codeine | 0.158 | 0.105 |
| Antimigraine | 0.016 | 0.004 |
| Topical analgesics | 0.174 | 0.117 |
| Topical NSAID | 0.152 | 0.103 |
| Lidocaine | 0.039 | 0.020 |
| Capsaicin | 0.003 | 0.002 |
| Pregabalin | 0.054 | 0.038 |
| Gabapentin | 0.010 | 0.007 |
| Amitriptyline | 0.033 | 0.017 |
| Paracetamol (inc. combinations) | 0.341 | 0.245 |
| Paracetamol (exc. opioid combinations) | 0.231 | 0.162 |
| Systemic NSAIDs | 0.328 | 0.245 |
| Non-selective NSAIDs | 0.305 | 0.226 |
| Coxib | 0.042 | 0.032 |
| All analgesics | 0.532 | 0.412 |
| **2016** |  |  |
| Opioids | 0.233 | 0.169 |
| Strong opioids^1^ | 0.112 | 0.083 |
| Long-acting opioids^2^ | 0.041 | 0.028 |
| Morphine | 0.008 | 0.007 |
| Hydromorphone | 0.000 | 0.000 |
| Oxycodone | 0.016 | 0.013 |
| Dihydrocodeine | 0.000 | 0.000 |
| Pethidine | 0.000 | 0.000 |
| Fentanyl | 0.006 | 0.003 |
| Buprenorphine | 0.016 | 0.007 |
| Tramadol | 0.080 | 0.061 |
| Meptazinol | 0.001 | 0.001 |
| Tapentadol | 0.009 | 0.006 |
| Codeine | 0.163 | 0.112 |
| Antimigraine | 0.017 | 0.004 |
| Topical analgesics | 0.186 | 0.125 |
| Topical NSAID | 0.158 | 0.108 |
| Lidocaine | 0.051 | 0.026 |
| Capsaicin | 0.004 | 0.002 |
| Pregabalin | 0.057 | 0.041 |
| Gabapentin | 0.011 | 0.008 |
| Amitriptyline | 0.034 | 0.018 |
| Paracetamol (inc. combinations) | 0.352 | 0.255 |
| Paracetamol (exc. opioid combinations) | 0.239 | 0.168 |
| Systemic NSAIDs | 0.330 | 0.246 |
| Non-selective NSAIDs | 0.307 | 0.227 |
| Coxib | 0.043 | 0.033 |
| All analgesics | 0.543 | 0.422 |
| **2017** |  |  |
| Opioids | 0.235 | 0.170 |
| Strong opioids^1^ | 0.113 | 0.084 |
| Long-acting opioids^2^ | 0.043 | 0.030 |
| Morphine | 0.009 | 0.008 |
| Hydromorphone | 0.000 | 0.000 |
| Oxycodone | 0.018 | 0.015 |
| Dihydrocodeine | 0.000 | 0.000 |
| Pethidine | 0.000 | 0.000 |
| Fentanyl | 0.006 | 0.003 |
| Buprenorphine | 0.017 | 0.008 |
| Tramadol | 0.077 | 0.058 |
| Meptazinol | 0.001 | 0.001 |
| Tapentadol | 0.011 | 0.008 |
| Codeine | 0.164 | 0.112 |
| Antimigraine | 0.017 | 0.004 |
| Topical analgesics | 0.193 | 0.131 |
| Topical NSAID | 0.166 | 0.116 |
| Lidocaine | 0.048 | 0.025 |
| Capsaicin | 0.004 | 0.003 |
| Pregabalin | 0.057 | 0.041 |
| Gabapentin | 0.013 | 0.009 |
| Amitriptyline | 0.037 | 0.020 |
| Paracetamol (inc. combinations) | 0.356 | 0.258 |
| Paracetamol (exc. opioid combinations) | 0.242 | 0.172 |
| Systemic NSAIDs | 0.321 | 0.236 |
| Non-selective NSAIDs | 0.297 | 0.217 |
| Coxib | 0.044 | 0.033 |
| All analgesics | 0.543 | 0.421 |
| **2018** |  |  |
| Opioids | 0.240 | 0.172 |
| Strong opioids^1^ | 0.113 | 0.084 |
| Long-acting opioids^2^ | 0.045 | 0.031 |
| Morphine | 0.009 | 0.008 |
| Hydromorphone | 0.000 | 0.000 |
| Oxycodone | 0.019 | 0.016 |
| Dihydrocodeine | 0.000 | 0.000 |
| Pethidine | 0.000 | 0.000 |
| Fentanyl | 0.006 | 0.003 |
| Buprenorphine | 0.018 | 0.008 |
| Tramadol | 0.073 | 0.056 |
| Meptazinol | 0.001 | 0.001 |
| Tapentadol | 0.013 | 0.009 |
| Codeine | 0.169 | 0.115 |
| Antimigraine | 0.017 | 0.004 |
| Topical analgesics | 0.176 | 0.122 |
| Topical NSAID | 0.172 | 0.119 |
| Lidocaine | 0.003 | 0.001 |
| Capsaicin | 0.006 | 0.003 |
| Pregabalin | 0.055 | 0.041 |
| Gabapentin | 0.014 | 0.010 |
| Amitriptyline | 0.039 | 0.021 |
| Paracetamol (inc. combinations) | 0.367 | 0.267 |
| Paracetamol (exc. opioid combinations) | 0.254 | 0.181 |
| Systemic NSAIDs | 0.319 | 0.230 |
| Non-selective NSAIDs | 0.296 | 0.211 |
| Coxib | 0.043 | 0.032 |
| All analgesics | 0.546 | 0.420 |
| **2019** |  |  |
| Opioids | 0.241 | 0.173 |
| Strong opioids^1^ | 0.112 | 0.083 |
| Long-acting opioids^2^ | 0.045 | 0.031 |
| Morphine | 0.010 | 0.009 |
| Hydromorphone | 0.000 | 0.000 |
| Oxycodone | 0.020 | 0.017 |
| Dihydrocodeine | 0.000 | 0.000 |
| Pethidine | 0.000 | 0.000 |
| Fentanyl | 0.006 | 0.003 |
| Buprenorphine | 0.018 | 0.008 |
| Tramadol | 0.070 | 0.053 |
| Meptazinol | 0.001 | 0.001 |
| Tapentadol | 0.014 | 0.010 |
| Codeine | 0.172 | 0.117 |
| Antimigraine | 0.016 | 0.004 |
| Topical analgesics | 0.177 | 0.122 |
| Topical NSAID | 0.174 | 0.120 |
| Lidocaine | 0.003 | 0.001 |
| Capsaicin | 0.004 | 0.003 |
| Pregabalin | 0.053 | 0.039 |
| Gabapentin | 0.015 | 0.010 |
| Amitriptyline | 0.041 | 0.022 |
| Paracetamol (inc. combinations) | 0.371 | 0.270 |
| Paracetamol (exc. opioid combinations) | 0.259 | 0.184 |
| Systemic NSAIDs | 0.310 | 0.218 |
| Non-selective NSAIDs | 0.288 | 0.200 |
| Coxib | 0.043 | 0.031 |
| All analgesics | 0.539 | 0.412 |
| **2020** |  |  |
| Opioids | 0.226 | 0.162 |
| Strong opioids^1^ | 0.105 | 0.079 |
| Long-acting opioids^2^ | 0.044 | 0.031 |
| Morphine | 0.011 | 0.010 |
| Hydromorphone | 0.000 | 0.000 |
| Oxycodone | 0.020 | 0.017 |
| Dihydrocodeine | 0.000 | 0.000 |
| Pethidine | 0.000 | 0.000 |
| Fentanyl | 0.005 | 0.003 |
| Buprenorphine | 0.017 | 0.008 |
| Tramadol | 0.062 | 0.047 |
| Meptazinol | 0.001 | 0.001 |
| Tapentadol | 0.015 | 0.011 |
| Codeine | 0.161 | 0.109 |
| Antimigraine | 0.014 | 0.004 |
| Topical analgesics | 0.159 | 0.108 |
| Topical NSAID | 0.156 | 0.107 |
| Lidocaine | 0.003 | 0.001 |
| Capsaicin | 0.002 | 0.001 |
| Pregabalin | 0.048 | 0.036 |
| Gabapentin | 0.015 | 0.011 |
| Amitriptyline | 0.041 | 0.021 |
| Paracetamol (inc. combinations) | 0.343 | 0.247 |
| Paracetamol (exc. opioid combinations) | 0.238 | 0.168 |
| Systemic NSAIDs | 0.272 | 0.188 |
| Non-selective NSAIDs | 0.251 | 0.171 |
| Coxib | 0.038 | 0.028 |
| All analgesics | 0.490 | 0.369 |
| **2021** |  |  |
| Opioids | 0.238 | 0.172 |
| Strong opioids^1^ | 0.110 | 0.083 |
| Long-acting opioids^2^ | 0.046 | 0.032 |
| Morphine | 0.011 | 0.010 |
| Hydromorphone | 0.000 | 0.000 |
| Oxycodone | 0.023 | 0.019 |
| Dihydrocodeine | 0.000 | 0.000 |
| Pethidine | 0.000 | 0.000 |
| Fentanyl | 0.005 | 0.003 |
| Buprenorphine | 0.019 | 0.009 |
| Tramadol | 0.063 | 0.048 |
| Meptazinol | 0.001 | 0.001 |
| Tapentadol | 0.016 | 0.012 |
| Codeine | 0.171 | 0.117 |
| Antimigraine | 0.016 | 0.004 |
| Topical analgesics | 0.172 | 0.117 |
| Topical NSAID | 0.169 | 0.116 |
| Lidocaine | 0.002 | 0.001 |
| Capsaicin | 0.003 | 0.002 |
| Pregabalin | 0.051 | 0.038 |
| Gabapentin | 0.018 | 0.012 |
| Amitriptyline | 0.044 | 0.023 |
| Paracetamol (inc. combinations) | 0.356 | 0.258 |
| Paracetamol (exc. opioid combinations) | 0.244 | 0.172 |
| Systemic NSAIDs | 0.288 | 0.198 |
| Non-selective NSAIDs | 0.266 | 0.180 |
| Coxib | 0.040 | 0.029 |
| All analgesics | 0.512 | 0.387 |
| **2022** |  |  |
| Opioids | 0.237 | 0.173 |
| Strong opioids^1^ | 0.106 | 0.080 |
| Long-acting opioids^2^ | 0.046 | 0.032 |
| Morphine | 0.010 | 0.010 |
| Hydromorphone | 0.000 | 0.000 |
| Oxycodone | 0.022 | 0.019 |
| Dihydrocodeine | 0.000 | 0.000 |
| Pethidine | 0.000 | 0.000 |
| Fentanyl | 0.005 | 0.003 |
| Buprenorphine | 0.018 | 0.009 |
| Tramadol | 0.060 | 0.046 |
| Meptazinol | 0.001 | 0.000 |
| Tapentadol | 0.016 | 0.011 |
| Codeine | 0.173 | 0.121 |
| Antimigraine | 0.016 | 0.004 |
| Topical analgesics | 0.173 | 0.120 |
| Topical NSAID | 0.171 | 0.118 |
| Lidocaine | 0.002 | 0.001 |
| Capsaicin | 0.004 | 0.002 |
| Pregabalin | 0.050 | 0.038 |
| Gabapentin | 0.019 | 0.013 |
| Amitriptyline | 0.045 | 0.023 |
| Paracetamol (inc. combinations) | 0.363 | 0.268 |
| Paracetamol (exc. opioid combinations) | 0.252 | 0.180 |
| Systemic NSAIDs | 0.291 | 0.200 |
| Non-selective NSAIDs | 0.270 | 0.183 |
| Coxib | 0.039 | 0.028 |
| All analgesics | 0.518 | 0.395 |

**Supplementary table 8. Absolute numbers of individuals with any use, initiations (90 days), discontinuations (90 days), and chronic use (90 days) over time.**

|  | **2014** | **2015** | **2016** | **2017** | **2018** | **2019** | **2020** | **2021** | **2022** |
| --- | --- | --- | --- | --- | --- | --- | --- | --- | --- |
| **Receipt of any dispensing** |  |  |  |  |  |  |  |  |  |
| **Opioids** | 348,434 | 340,031 | 341,810 | 330,013 | 327,095 | 324,319 | 312,204 | 321,406 | 326,223 |
| *Strong opioids^1^* | 164,489 | 164,811 | 165,012 | 159,372 | 154,940 | 152,893 | 147,385 | 151,432 | 147,428 |
| *Long-acting opioids^2^* | 53,061 | 53,780 | 58,933 | 59,563 | 59,472 | 60,231 | 60,222 | 61,809 | 62,735 |
| *Morphine* | 12,381 | 12,144 | 13,470 | 12,879 | 14,085 | 13,899 | 15,848 | 15,452 | 15,684 |
| *Hydromorphone* | <1,500 | <1,500 | <1,500 | <1,500 | <1,500 | <1,500 | <1,500 | <1,500 | <1,500 |
| *Oxycodone* | 21,224 | 22,553 | 25,257 | 27,367 | 28,171 | 29,343 | 28,526 | 32,450 | 32,936 |
| *Dihydrocodeine* | 19,456 | 6,939 | <1,500 | <1,500 | <1,500 | <1,500 | <1,500 | <1,500 | <1,500 |
| *Pethidine* | <1,500 | <1,500 | <1,500 | <1,500 | <1,500 | <1,500 | 0 | 0 | 0 |
| *Fentanyl* | 8,844 | 8,674 | 8,419 | 8,049 | 7,825 | 7,722 | 6,339 | 7,726 | 6,274 |
| *Buprenorphine* | 19,456 | 19,083 | 20,206 | 20,928 | 21,911 | 21,621 | 20,602 | 21,633 | 21,957 |
| *Tramadol* | 129,115 | 124,909 | 119,549 | 109,468 | 101,728 | 95,751 | 87,163 | 86,532 | 84,692 |
| *Meptazinol* | 1,769 | 1,735 | 1,684 | 1,610 | 1,565 | 1,544 | 1,585 | 1,545 | 1,568 |
| *Tapentadol* | 5,306 | 8,674 | 13,470 | 16,098 | 17,216 | 18,532 | 20,602 | 21,633 | 21,957 |
| *Codeine* | 226,394 | 230,735 | 234,047 | 225,375 | 225,367 | 227,023 | 217,116 | 225,602 | 233,688 |
| **Antimigraine** | 19,456 | 19,083 | 18,522 | 17,708 | 17,216 | 15,444 | 15,848 | 15,452 | 15,684 |
| **Topical analgesics** | 247,618 | 255,023 | 266,039 | 264,010 | 236,322 | 234,745 | 213,947 | 227,148 | 233,688 |
| *Topical NSAID* | 222,856 | 223,796 | 227,312 | 230,204 | 231,627 | 230,112 | 210,777 | 224,057 | 230,552 |
| *Lidocaine* | 40,680 | 52,046 | 67,352 | 59,563 | 3,130 | 3,089 | 3,170 | 3,090 | 3,137 |
| *Capsaicin* | 5,306 | 5,205 | 5,051 | 6,439 | 7,825 | 4,633 | 3,170 | 4,636 | 4,705 |
| **Pregabalin** | 77,823 | 79,803 | 82,506 | 80,491 | 75,122 | 71,041 | 68,146 | 69,535 | 70,577 |
| **Gabapentin** | 14,150 | 15,614 | 16,838 | 17,708 | 18,781 | 20,077 | 20,602 | 23,178 | 25,094 |
| **Amitriptyline** | 42,449 | 43,371 | 45,462 | 46,685 | 48,517 | 49,420 | 50,713 | 54,083 | 54,893 |
| **Paracetamol (inc. combinations)** | 525,304 | 513,516 | 516,924 | 499,044 | 502,381 | 501,922 | 473,852 | 480,564 | 501,881 |
| *Paracetamol (exc. opioid combinations)* | 348,434 | 345,236 | 346,861 | 338,062 | 344,311 | 345,940 | 326,467 | 326,042 | 343,475 |
| **Systemic NSAIDs** | 519,998 | 501,373 | 489,983 | 453,969 | 435,084 | 413,892 | 369,256 | 381,670 | 392,095 |
| *Non-selective NSAIDs* | 482,855 | 464,941 | 452,940 | 418,553 | 402,218 | 381,460 | 339,145 | 350,765 | 362,296 |
| *Coxib* | 65,442 | 65,924 | 63,984 | 62,783 | 59,472 | 57,142 | 52,298 | 54,083 | 53,325 |
| **All analgesics** | 854,282 | 825,790 | 818,323 | 782,373 | 763,744 | 741,300 | 687,799 | 703,076 | 726,159 |
| **Initiations** |  |  |  |  |  |  |  |  |  |
| **Opioids** | 146,802 | 220,326 | 230,680 | 225,375 | 222,237 | 220,845 | 209,192 | 214,786 | 219,573 |
| *Strong opioids^1^* | 74,285 | 109,296 | 111,130 | 107,858 | 103,293 | 100,384 | 96,672 | 98,894 | 95,671 |
| *Long-acting opioids^2^* | 21,224 | 32,962 | 35,360 | 35,416 | 34,431 | 33,976 | 34,865 | 35,540 | 36,073 |
| *Morphine* | 7,075 | 10,409 | 10,103 | 11,269 | 10,955 | 12,355 | 14,263 | 12,362 | 12,547 |
| *Hydromorphone* | <1,500 | <1,500 | <1,500 | <1,500 | <1,500 | <1,500 | <1,500 | <1,500 | <1,500 |
| *Oxycodone* | 10,612 | 15,614 | 18,522 | 19,318 | 20,346 | 21,621 | 20,602 | 24,724 | 23,526 |
| *Dihydrocodeine* | 8,844 | 3,470 | <1,500 | <1,500 | <1,500 | <1,500 | <1,500 | <1,500 | <1,500 |
| *Pethidine* | <1,500 | <1,500 | <1,500 | <1,500 | <1,500 | <1,500 | 0 | 0 | 0 |
| *Fentanyl* | 3,537 | 3,470 | 3,368 | 3,220 | 3,130 | 3,089 | 3,170 | 3,090 | 3,137 |
| *Buprenorphine* | 8,844 | 12,144 | 11,787 | 12,879 | 12,520 | 12,355 | 11,094 | 12,362 | 12,547 |
| *Tramadol* | 60,136 | 88,478 | 87,557 | 78,881 | 73,557 | 67,952 | 61,807 | 60,264 | 58,030 |
| *Meptazinol* | <1,500 | <1,500 | 1,684 | 1,610 | 1,565 | 1,544 | 1,585 | 1,545 | <1,500 |
| *Tapentadol* | 3,537 | 6,939 | 10,103 | 12,879 | 14,085 | 13,899 | 15,848 | 15,452 | 15,684 |
| *Codeine* | 100,816 | 163,076 | 171,747 | 167,421 | 169,025 | 169,881 | 160,064 | 166,884 | 172,522 |
| **Antimigraine** | 7,075 | 12,144 | 11,787 | 11,269 | 10,955 | 10,811 | 9,509 | 10,817 | 10,979 |
| **Topical analgesics** | 125,578 | 189,099 | 203,739 | 202,837 | 186,241 | 185,325 | 164,818 | 171,520 | 177,227 |
| *Topical NSAID* | 111,428 | 168,281 | 176,798 | 183,519 | 183,111 | 182,236 | 163,233 | 169,974 | 175,658 |
| *Lidocaine* | 22,993 | 41,636 | 52,198 | 43,465 | 3,130 | 1,544 | 1,585 | 1,545 | 1,568 |
| *Capsaicin* | 1,769 | 3,470 | 3,368 | 4,829 | 6,260 | 4,633 | 3,170 | 3,090 | 4,705 |
| **Pregabalin** | 28,299 | 41,636 | 42,095 | 38,636 | 34,431 | 32,432 | 28,526 | 29,359 | 29,799 |
| **Gabapentin** | 5,306 | 8,674 | 8,419 | 9,659 | 10,955 | 10,811 | 11,094 | 12,362 | 14,115 |
| **Amitriptyline** | 14,150 | 22,553 | 21,889 | 22,537 | 23,476 | 24,710 | 23,772 | 24,724 | 25,094 |
| **Paracetamol (inc. combinations)** | 215,781 | 322,683 | 341,810 | 336,452 | 338,051 | 335,129 | 309,034 | 307,499 | 319,949 |
| *Paracetamol (exc. opioid combinations)* | 152,108 | 229,001 | 240,782 | 238,253 | 242,583 | 242,467 | 221,871 | 216,331 | 227,415 |
| **Systemic NSAIDs** | 217,550 | 338,296 | 360,331 | 347,721 | 338,051 | 327,407 | 294,771 | 304,409 | 310,539 |
| *Non-selective NSAIDs* | 201,632 | 315,743 | 336,758 | 323,574 | 316,140 | 307,330 | 275,753 | 284,321 | 291,718 |
| *Coxib* | 33,605 | 48,576 | 50,514 | 49,904 | 46,951 | 46,331 | 41,205 | 41,721 | 42,346 |
| **All analgesics** | 290,067 | 449,327 | 481,565 | 470,067 | 460,124 | 447,868 | 404,121 | 406,393 | 417,189 |
| **Discontinuation** |  |  |  |  |  |  |  |  |  |
| **Opioids** | 123,809 | 182,160 | 190,268 | 186,739 | 183,111 | 182,236 | 169,573 | 174,610 | 180,364 |
| *Strong opioids^1^* | 63,673 | 90,212 | 94,292 | 91,760 | 87,643 | 84,941 | 77,655 | 80,352 | 81,556 |
| *Long-acting opioids^2^* | 15,918 | 24,288 | 26,941 | 27,367 | 26,606 | 27,799 | 25,357 | 27,814 | 28,231 |
| *Morphine* | 3,537 | 3,470 | 3,368 | 4,829 | 4,695 | 4,633 | 4,754 | 4,636 | 4,705 |
| *Hydromorphone* | <1,500 | <1,500 | <1,500 | <1,500 | <1,500 | <1,500 | <1,500 | <1,500 | <1,500 |
| *Oxycodone* | 8,844 | 12,144 | 15,154 | 16,098 | 17,216 | 16,988 | 15,848 | 20,088 | 20,389 |
| *Dihydrocodeine* | 8,844 | 10,409 | 1,684 | <1,500 | <1,500 | <1,500 | <1,500 | <1,500 | <1,500 |
| *Pethidine* | <1,500 | <1,500 | <1,500 | <1,500 | <1,500 | <1,500 | 0 | 0 | 0 |
| *Fentanyl* | 1,769 | 3,470 | 3,368 | 3,220 | 3,130 | 3,089 | 1,585 | 1,545 | 1,568 |
| *Buprenorphine* | 5,306 | 8,674 | 8,419 | 8,049 | 9,390 | 9,266 | 7,924 | 9,271 | 9,410 |
| *Tramadol* | 56,598 | 81,538 | 82,506 | 75,662 | 70,427 | 64,864 | 58,637 | 57,173 | 54,893 |
| *Meptazinol* | <1,500 | <1,500 | <1,500 | 1,610 | 1,565 | 1,544 | 1,585 | 1,545 | <1,500 |
| *Tapentadol* | 1,769 | 5,205 | 8,419 | 9,659 | 10,955 | 12,355 | 12,678 | 15,452 | 14,115 |
| *Codeine* | 90,204 | 138,788 | 148,174 | 146,494 | 145,550 | 146,716 | 137,877 | 143,706 | 150,564 |
| **Antimigraine** | 5,306 | 8,674 | 10,103 | 9,659 | 9,390 | 9,266 | 7,924 | 7,726 | 7,842 |
| **Topical analgesics** | 109,659 | 163,076 | 176,798 | 181,910 | 172,155 | 163,704 | 145,801 | 148,341 | 156,838 |
| *Topical NSAID* | 100,816 | 149,197 | 158,276 | 157,762 | 161,200 | 160,615 | 144,216 | 146,796 | 155,270 |
| *Lidocaine* | 17,687 | 31,227 | 40,411 | 49,904 | 20,346 | 3,089 | 1,585 | 1,545 | 1,568 |
| *Capsaicin* | 1,769 | 3,470 | 3,368 | 4,829 | 6,260 | 6,177 | 3,170 | 3,090 | 3,137 |
| **Pregabalin** | 22,993 | 32,962 | 35,360 | 33,806 | 31,301 | 27,799 | 23,772 | 24,724 | 25,094 |
| **Gabapentin** | 3,537 | 6,939 | 6,735 | 8,049 | 9,390 | 9,266 | 9,509 | 10,817 | 10,979 |
| **Amitriptyline** | 12,381 | 17,349 | 20,206 | 19,318 | 18,781 | 20,077 | 19,017 | 20,088 | 20,389 |
| **Paracetamol (inc. combinations)** | 175,101 | 248,084 | 260,988 | 255,961 | 250,408 | 248,644 | 229,795 | 228,693 | 236,825 |
| *Paracetamol (exc. opioid combinations)* | 127,346 | 183,894 | 193,636 | 189,959 | 187,806 | 188,414 | 174,327 | 168,429 | 175,658 |
| **Systemic NSAIDs** | 180,407 | 253,289 | 267,723 | 260,791 | 253,538 | 253,277 | 229,795 | 236,419 | 243,099 |
| *Non-selective NSAIDs* | 168,027 | 239,410 | 252,569 | 244,693 | 239,452 | 240,922 | 217,116 | 222,512 | 228,983 |
| *Coxib* | 31,837 | 45,106 | 47,146 | 45,075 | 45,386 | 43,242 | 38,035 | 40,176 | 39,209 |
| **All analgesics** | 166,258 | 196,038 | 200,371 | 198,008 | 195,631 | 194,591 | 177,496 | 179,246 | 186,637 |
| **Chronic use** |  |  |  |  |  |  |  |  |  |
| **Opioids** | 97,279 | 105,826 | 107,763 | 106,248 | 107,988 | 111,195 | 115,690 | 118,982 | 117,628 |
| *Strong opioids^1^* | 53,061 | 57,250 | 60,617 | 61,173 | 61,037 | 61,775 | 61,807 | 63,354 | 61,167 |
| *Long-acting opioids^2^* | 24,762 | 27,758 | 30,308 | 30,587 | 31,301 | 30,887 | 31,696 | 30,904 | 31,368 |
| *Morphine* | 3,537 | 3,470 | 3,368 | 3,220 | 3,130 | 4,633 | 4,754 | 4,636 | 4,705 |
| *Hydromorphone* | <1,500 | <1,500 | <1,500 | <1,500 | <1,500 | <1,500 | <1,500 | <1,500 | <1,500 |
| *Oxycodone* | 8,844 | 8,674 | 10,103 | 9,659 | 10,955 | 10,811 | 11,094 | 10,817 | 10,979 |
| *Dihydrocodeine* | 3,537 | 1,735 | <1,500 | <1,500 | <1,500 | <1,500 | <1,500 | <1,500 | 0 |
| *Pethidine* | <1,500 | <1,500 | <1,500 | <1,500 | <1,500 | <1,500 | 0 | 0 | 0 |
| *Fentanyl* | 3,537 | 5,205 | 5,051 | 4,829 | 4,695 | 4,633 | 4,754 | 4,636 | 4,705 |
| *Buprenorphine* | 8,844 | 10,409 | 11,787 | 11,269 | 12,520 | 12,355 | 12,678 | 12,362 | 12,547 |
| *Tramadol* | 28,299 | 31,227 | 30,308 | 28,977 | 28,171 | 27,799 | 26,941 | 27,814 | 26,662 |
| *Meptazinol* | <1,500 | <1,500 | <1,500 | <1,500 | <1,500 | <1,500 | <1,500 | <1,500 | <1,500 |
| *Tapentadol* | 1,769 | 3,470 | 3,368 | 4,829 | 6,260 | 6,177 | 6,339 | 7,726 | 6,274 |
| *Codeine* | 40,680 | 46,841 | 48,830 | 48,295 | 50,082 | 50,964 | 55,468 | 58,718 | 59,598 |
| **Antimigraine** | 5,306 | 5,205 | 5,051 | 4,829 | 6,260 | 4,633 | 4,754 | 4,636 | 6,274 |
| **Topical analgesics** | 40,680 | 50,311 | 55,565 | 56,344 | 43,821 | 46,331 | 52,298 | 58,718 | 59,598 |
| *Topical NSAID* | 30,068 | 36,432 | 37,043 | 38,636 | 42,256 | 44,787 | 50,713 | 55,628 | 58,030 |
| *Lidocaine* | 8,844 | 13,879 | 16,838 | 19,318 | 1,565 | 1,544 | 1,585 | 1,545 | 1,568 |
| *Capsaicin* | <1,500 | <1,500 | <1,500 | <1,500 | 1,565 | 1,544 | <1,500 | <1,500 | <1,500 |
| **Pregabalin** | 38,911 | 43,371 | 47,146 | 46,685 | 45,386 | 44,787 | 44,374 | 44,811 | 43,915 |
| **Gabapentin** | 7,075 | 6,939 | 8,419 | 8,049 | 9,390 | 9,266 | 11,094 | 12,362 | 12,547 |
| **Amitriptyline** | 28,299 | 29,493 | 31,992 | 32,196 | 32,866 | 35,521 | 36,450 | 38,631 | 39,209 |
| **Paracetamol (inc. combinations)** | 132,653 | 147,463 | 151,541 | 151,323 | 158,070 | 163,704 | 177,496 | 186,972 | 191,342 |
| *Paracetamol (exc. opioid combinations)* | 74,285 | 86,743 | 89,241 | 90,150 | 93,903 | 98,840 | 110,935 | 117,437 | 120,765 |
| **Systemic NSAIDs** | 56,598 | 62,455 | 58,933 | 56,344 | 54,777 | 54,053 | 57,052 | 58,718 | 56,462 |
| *Non-selective NSAIDs* | 44,218 | 48,576 | 45,462 | 43,465 | 42,256 | 41,698 | 42,789 | 44,811 | 43,915 |
| *Coxib* | 10,612 | 12,144 | 11,787 | 11,269 | 10,955 | 10,811 | 11,094 | 12,362 | 10,979 |
| **All analgesics** | 314,829 | 334,827 | 340,126 | 338,062 | 336,486 | 339,762 | 347,069 | 360,037 | 365,432 |

**Supplementary table 9. Prevalence (%) of high-risk dispensings in the GMS population**

|  | **2014** | **2015** | **2016** | **2017** | **2018** | **2019** | **2020** | **2021** | **2022** |
| --- | --- | --- | --- | --- | --- | --- | --- | --- | --- |
| **Non-selective NSAIDs prescribed with medications increasing the risk of bleeding** | | | | | | | | | |
| Prevalence of nsNSAIDs with antiplatelets* | 3.9 | 3.6 | 3.6 | 3.5 | 3.6 | 3.5 | 3.0 | 3.1 | 3.2 |
| *Percentage of all nsNSAID dispensings with antiplatelets* | 14.1 | 13.4 | 13.3 | 13.6 | 13.8 | 14.3 | 14.2 | 13.8 | 13.7 |
| Prevalence nsNSAIDs with oral anticoagulants* | 0.6 | 0.7 | 0.8 | 0.9 | 0.9 | 1.00 | 0.9 | 1.0 | 1.1 |
| *Percentage of all nsNSAID dispensings with oral anticoagulants* | 2.1 | 2.5 | 2.9 | 3.3 | 3.7 | 4.1 | 4.3 | 4.6 | 4.9 |
| Prevalence of nsNSAIDs with oral corticosteroids* | 0.5 | 0.5 | 0.6 | 0.6 | 0.7 | 0.7 | 0.6 | 0.7 | 0.7 |
| *Percentage of all nsNSAID dispensings with oral corticosteroids* | 1.7 | 2.0 | 2.2 | 2.4 | 2.6 | 2.9 | 2.9 | 3.1 | 3.1 |
| Prevalence of nsNSAIDs with SSRIs* | 3.0 | 3.3 | 3.6 | 3.7 | 3.9 | 4.0 | 3.7 | 4.1 | 4.2 |
| *Percentage of all nsNSAID dispensings with SSRIs* | 10.9 | 12.5 | 13.4 | 14.4 | 15.2 | 16.4 | 17.3 | 18.0 | 18.2 |
| **Non-selective NSAIDs prescribed with medications increasing the risk of acute kidney injury** | | | | | | | | | |
| Prevalence of nsNSAIDs with ACEi/ARB and diuretics* | 1.9 | 1.8 | 1.8 | 1.8 | 1.8 | 1.8 | 1.6 | 1.7 | 1.7 |
| *Percentage of all nsNSAID dispensings* *with ACEi/ARB and diuretics* | 7.1 | 6.9 | 6.8 | 7.0 | 7.1 | 7.4 | 7.4 | 7.3 | 7.3 |
| Prevalence of nsNSAIDs with ACEi/ARB and metformin in people aged ≥65 years* | 1.7 | 1.2 | 1.0 | 0.9 | 0.8 | 0.8 | 0.8 | 0.8 | 0.9 |
| *Percentage of all nsNSAID dispensings with ACEi/ARB and metformin to people aged ≥65 years* | 9.9 | 7.3 | 6.2 | 5.5 | 5.7 | 5.6 | 5.8 | 5.7 | 6.1 |
| **Non-selective NSAIDs prescribed with medications increasing the risk of seizures** | | | | | | | | | |
| Prevalence of nsNSAIDs with lithium* | 0.09 | 0.09 | 0.10 | 0.10 | 0.09 | 0.10 | 0.08 | 0.09 | 0.08 |
| *Percentage of all nsNSAID dispensings with lithium* | 0.3 | 0.4 | 0.4 | 0.4 | 0.4 | 0.4 | 0.4 | 0.4 | 0.4 |
| **Opioids prescribed with medications increasing the risk of falls, fractures and delirium** | | | | | | | | | |
| Prevalence of opioids with two or more other sedating or anticholinergic drugs in people aged ≥65 years* | 15.4 | 15.3 | 15.2 | 14.8 | 14.8 | 14.6 | 14.2 | 14.1 | 14.0 |
| *Percentage of all opioid dispensings with two or more other sedating or anticholinergic drugs to people aged ≥65 years* | 72.0 | 69.8 | 66.9 | 66.1 | 71.6 | 71.8 | 70.7 | 67.7 | 65.0 |
| **Dispensings at a dose increasing the risk of dependency** | | | | | | | | | |
| Prevalence of opioids dispensed at dose equivalent to >90 mg morphine per day* | 1.2 | 1.2 | 1.4 | 1.4 | 1.5 | 1.5 | 1.4 | 1.5 | 1.4 |
| *Percentage of all opioid dispensings at dose equivalent to >90 mg morphine per day* | 5.9 | 6.3 | 6.6 | 6.9 | 6.9 | 7.0 | 7.3 | 7.0 | 6.7 |
| Prevalence of pregabalin dispensed at a dose >800 mg per day* | 0.31 | 0.35 | 0.37 | 0.38 | 0.38 | 0.38 | 0.35 | 0.36 | 0.36 |
| *Percentage of all pregabalin dispensings at a dose >800 mg per day* | 7.0 | 7.5 | 7.5 | 7.7 | 7.9 | 8.1 | 8.3 | 8.0 | 8.1 |
| Prevalence of gabapentin dispensed at a dose >4800 mg per day* | 0.03 | 0.03 | 0.03 | 0.03 | 0.03 | 0.04 | 0.04 | 0.04 | 0.04 |
| *Percentage of all gabapentin dispensings at a dose >4800 mg per day* | 3.7 | 3.6 | 3.5 | 3.0 | 2.8 | 2.9 | 2.8 | 2.5 | 2.5 |

* Prevalence figures are number of individuals with the specified high-risk dispensing as a percentage of all GMS eligible individual
